# City-wide school-located influenza vaccination: a retrospective cohort study

**DOI:** 10.1101/2021.05.03.21256546

**Authors:** Jade Benjamin-Chung, Benjamin F. Arnold, Kunal Mishra, Chris J. Kennedy, Anna Nguyen, Nolan N. Pokpongkiat, Stephanie Djajadi, Anmol Seth, Nicola P. Klein, Alan E. Hubbard, Arthur Reingold, John M. Colford

## Abstract

**Background:** We measured the effectiveness of a city-wide school-located influenza vaccination (SLIV) program implemented in over 102 elementary schools in Oakland, California.

**Methods:** We conducted a retrospective cohort study among Kaiser Permanente Northern California (KPNC) members of all ages residing in either the intervention or a multivariate-matched comparison site from September 2011 - August 2017. Outcomes included medically attended acute respiratory illness (MAARI), influenza hospitalization, and Oseltamivir prescriptions. We estimated difference-in-differences (DIDs) in 2014-15, 2015-16, and 2016-17 using generalized linear models and adjusted for race, ethnicity, age, sex, health plan, and language.

**Results:** Pre-intervention member characteristics were similar between sites. Among school-aged children, SLIV was associated with lower Oseltamivir prescriptions per 1,000 (DIDs: −3.5 (95% CI −5.5, −1.5) in 2015-16; −4.0 (95% CI −6.5, −1.6) in 2016-17) but not with other outcomes. SLIV was associated with lower MAARI per 1,000 in adults 65+ years (2014-15: −13.2, 95% CI −23.2, −3.2; 2015-16: −21.5, 95% CI −31.1, −11.9; 2016-17: −13.0, 95% CI −23.2, −2.9). There were few significant associations with other outcomes among adults.

**Conclusions:** A city-wide SLIV intervention was associated with lower Oseltamivir prescriptions in school-aged children and lower MAARI among people over 65 years, suggesting possible indirect effects of SLIV among older adults.

## Introduction

In the United States, seasonal influenza has been responsible for 140,000 – 590,000 hospitalizations and 12,000 – 61,000 deaths annually since 2010 [1]. Children are responsible for the majority of influenza transmission, and mathematical models suggest that vaccinating 50% – 70% of school-aged children for influenza can produce herd immunity [2].

School-located influenza vaccination (SLIV) interventions may increase vaccine coverage among schoolchildren and reduce influenza transmission community-wide [3]. SLIV is associated with higher influenza vaccination coverage [4–13] and lower medically attended acute respiratory illness (MAARI) [14], influenza-like illness [4] and laboratory-confirmed influenza [14,15] in schoolchildren. Some studies have reported indirect effects of SLIV among non-school aged individuals, while others found none [7,8,14,16–20]. With the exception of one study [4], prior SLIV evaluations have measured health outcomes using observational designs that did not rigorously account for systematic differences between intervention and comparison sites prior to intervention [5–8,13–20].

We previously reported the findings from a matched cohort study of a city-wide SLIV program called Shoo the Flu that was implemented in a diverse, primarily low-income population in Oakland, California [21]. The initial evaluation found higher vaccination coverage and lower influenza hospitalization among non-elementary school aged individuals in the intervention site.

Here, we report the results of a retrospective cohort study to measure associations between Shoo the Flu program on additional outcomes, including MAARI and Oseltamivir prescriptions, among Kaiser Permanente Northern California (KPNC) members residing within either the intervention or a matched comparison area. Using data from 2011 to 2017, we estimated associations with SLIV among school-aged individuals and assessed potential indirect effects in other age groups.

## Methods

### SLIV intervention

This study evaluated the Shoo the Flu intervention (www.shootheflu.org), which has delivered free influenza vaccinations to schools in Oakland, California since 2014. The intervention was delivered to children in all public and charter elementary schools in Oakland Unified School District (OUSD, the “intervention district”) and offered to all other charter and private pre-schools and elementary schools in Oakland. OUSD has a diverse population of over 26,000 elementary school students, and >70% are low-income. From 2014-2017, Shoo the Flu vaccinated 7,502 – 10,106 students annually (22 – 28% of eligible students) in 102-138 schools. Each influenza season, 23-24% of intervention participants reported KPNC health plan membership. Additional intervention details are in the Supplement 1.

### Study setting and population

KPNC is an integrated healthcare system that delivers care at 46 medical clinics and 21 hospitals operated by KPNC to approximately 4 million members. Members comprise at least 30% of the population and are representative of the race, ethnicity, and socioeconomic distribution of Northern California, although very low-income individuals are under-represented. Health care visits, diagnoses, prescriptions, immunizations, and laboratory results are captured in KPNC’s electronic medical record. Vaccines are offered free of charge to members, and the date, injection site, and vaccine brand, lot, and dose of each vaccination at KPNC are recorded. Whether to test patients for influenza A or B using polymerase chain reaction is at the discretion of KPNC clinicians.

All KPNC members who resided in the catchment areas of the intervention and comparison districts from September 1, 2011 - August 31, 2017 and had no more than a one-month gap in KPNC membership for each influenza season during the study period were included in this study.

### Vaccines

In 2014-15 and 2015-16, the intervention provided the live attenuated influenza vaccine (LAIV) to students [22,23]. Students with LAIV contraindications were offered the trivalent inactivated injectable influenza vaccine (IIV3), as were staff and teachers. Because LAIV effectiveness in children was low in 2014-15 and 2015-16 [24–26], the intervention offered IIV4 to all participants following the Advisory Committee on Immunization Practices’ recommendation to use IIV for all children in 2016-17 [25].

### Study design

We conducted a retrospective cohort study of KPNC health plan members who lived in the catchment areas of the intervention district and a comparison district (West Contra Costa Unified School District [WCCUSD]). We identified the comparison school district using a genetic multivariate matching algorithm [27] to pair-match public elementary schools in the intervention district and each candidate comparison district using pre-intervention school-level characteristics (additional details in Supplement 2); detailed methods are available elsewhere [21]. We selected WCCUSD as the comparison site because it had the smallest average generalized Mahalanobis distance between paired schools [27].

### Program data

KPNC electronic medical records did not include records for vaccinations administered at locations other than KPNC. We therefore estimated the number of vaccinations delivered by the program to KPNC members using data from the Shoo the Flu program, which tracks vaccination counts using the number of vaccination consent forms collected from the parents each year. Consent forms included information about insurance provider (e.g., KPNC or other provider), allowing us to estimate the number of children vaccinated by Shoo the Flu who were KPNC members.

### Population and school district data

We obtained demographic information about the general population in study sites from the U.S. 2010 Census using zip codes that overlapped with the intervention and comparison school districts. We also obtained data about school district populations from the California Department of Education for these zip codes for 2013.

### Outcomes

Outcomes included medically attended acute respiratory illness (MAARI) (see Supplement Table 1 for ICD-9 and ICD-10 codes), laboratory-confirmed influenza among tested individuals, influenza hospitalization, and filled Oseltamivir prescriptions. Individuals hospitalized with any of the following ICD-9-CM codes for otitis media and sinusitis (381–383, 461x), upper respiratory tract illness (79x, 460, 462–463, 465, 487.1), and lower respiratory tract illness (464x, 466x, 480x–487.0, 490x–496x, 510x–513x, 515x–516x, 518x, and 786.1) were classified as having an influenza hospitalization. We defined laboratory-confirmed influenza as a positive result from RT-PCR influenza diagnostic test.

### Definition of influenza season

We defined influenza season based on the percentage of medical visits for influenza-like illness in California using data from the California Department of Public Health. Influenza season started after two consecutive weeks in which the percentage of medical visits for influenza-like illness was greater than or equal to 2%; it ended after two consecutive weeks with a percentage under 2%.

### Statistical analysis

This study’s pre-analysis plan and replication scripts are available at https://osf.io/rtsf2/.

We defined the cumulative incidence as the proportion of individuals with at least one outcome event in each season. We restricted analyses to influenza season, when the intervention would be expected to affect influenza-related outcomes. Our primary pre-specified parameter was mean difference-in-differences (DIDs) that compared the difference in cumulative incidence during influenza season to that in three seasons prior to the intervention (2011-2014) in each study group (the “pre-intervention DID”). Pre-intervention monthly incidences of each outcome were consistent with the equal trends assumption (Supplement Figure 2). DIDs remove time-invariant confounding but may be subject to time-dependent confounding [28]. To account for this, we conducted a post-hoc alternative “pre-season DID” analysis that compared the incidence in each influenza season to that in the period immediately preceding each season (May – September). While the pre-season DID does not account for pre-intervention differences, it is less subject to time-dependent confounding than the pre-intervention DID.

Models adjusted for available potential confounders with at least 5% prevalence in each analysis; these included race, ethnicity, sex, mediCAL, subsidized KPNC health plan (proxy for low socio-economic status), and primary language spoken. Enrollment in mediCAL or a subsidized KPNC health plan varied by season; other variables were static. To minimize empirical positivity violations from sparse data [29], we fit models only if the number of outcome events per variable was ≥10 and only fit adjusted models if the number of observations within age, site, and outcome strata was ≥30 [30]. We estimated 95% confidence intervals using robust standard errors [31] to account for clustering at the household level. Additional minor deviations from the plan are noted in Supplement 3, and additional analysis details are in Supplement 4.

We stratified estimates by pre-specified age groups (0-4, 5-12, 13-17, 18-64, and 65+ years). Estimates among children 5-12 years represent “total effects” (including intervention participants and non-participants) and estimates in other age groups represent “indirect effects” among non-participants (Supplement Figure 1) [32]. Per our pre-analysis plan, we also stratified estimates by individual vaccination status.

We performed a sensitivity analysis using alternative influenza seasons with influenza-like-illness thresholds of 2.5% and 3% and the CDC influenza season definition (Week 40 to Week 20 of the following year).

We pre-specified two negative control analyses to detect residual confounding or selection bias [33,34]. First, we repeated our primary analysis with outcomes we did not expect SLIV to affect (medically attended diarrhea and medically attended gastrointestinal illness) [33,34]. We conducted a negative control time period analysis restricting to weeks outside influenza season.

### Ethical statement

This study was approved by the Committee for the Protection of Human Subjects at the University of California, Berkeley (Protocol # 2017-03-9741) and the KPNC Institutional Review Board (Protocol #CN-16-2825).

## Results

### Pre-intervention characteristics

The analyses included 175,628 to 269,266 individuals and 9,436,202 to 11,500,570 person-weeks of observations per calendar year from 2011 to 2017 (Supplement Table 2). During the influenza season, the number of person-weeks ranged from 3,069,633 to 6,801,780 per year.

Pre-intervention characteristics were similar among individuals in the intervention and comparison sites (Table 1). In the intervention site vs. the comparison site, there were fewer Asian (14.2% vs. 19.3%) and Hispanic (30.5% vs. 43.0%) individuals and more Black / African American (28.2% vs. 20.2%) and White individuals (35.2% vs. 29.5%). The proportion of individuals enrolled in MediCAL was lower in the intervention site vs. the comparison site (4.4% vs. 6.7%).

**Table 1.**
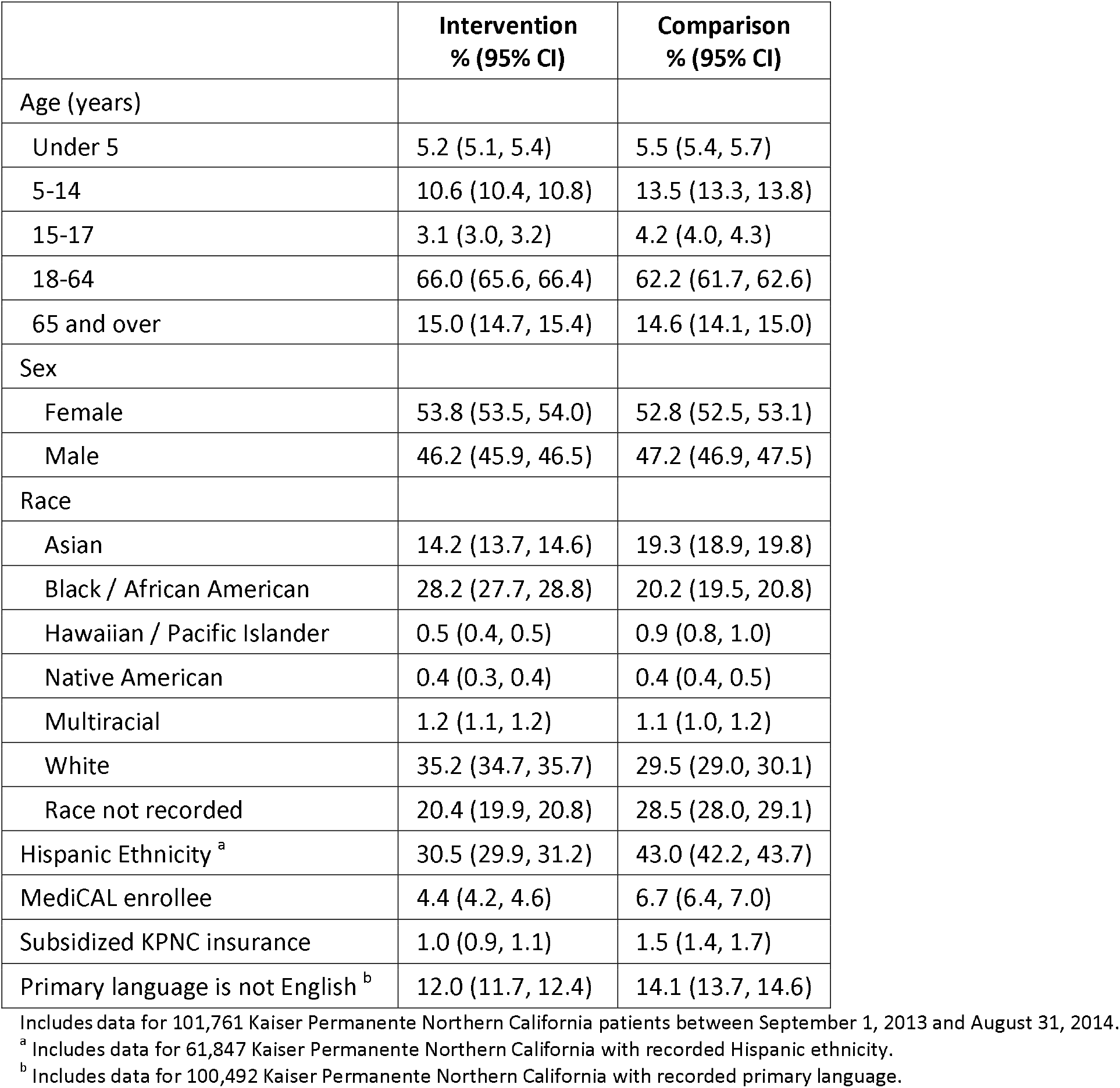
Pre-intervention characteristics of the study population in each site.

Characteristics were similar overall between the study population, general population (all individuals residing in zip codes overlapping with the study site) and school district population (Supplement Table 3). The percentage of individuals in the study population who were Hispanic was lower than in the general population (31-43% vs. 39-54%). The percentage of the study population whose primary language spoken was not English was also lower (12-14%) than in the general population (40-47%) and school district population (38-43%).

### Influenza vaccination

Among the study population, influenza vaccination coverage was lower in the intervention site than the comparison site across age groups and years. Prior to the intervention, vaccination coverage was also consistently lower in the intervention site than the comparison site (Supplement Table 4); this difference was larger during the intervention period for children 5-12 years (Supplement Figure 3). Including all vaccinations (administered by both KPNC and the SLIV intervention), coverage was 8-11% higher in the intervention site than the comparison site during the intervention period (Figure 1). Across all years, KPNC mostly administered IIV to elementary school aged children in the study (Supplement Figure 4).

**Figure 1.**
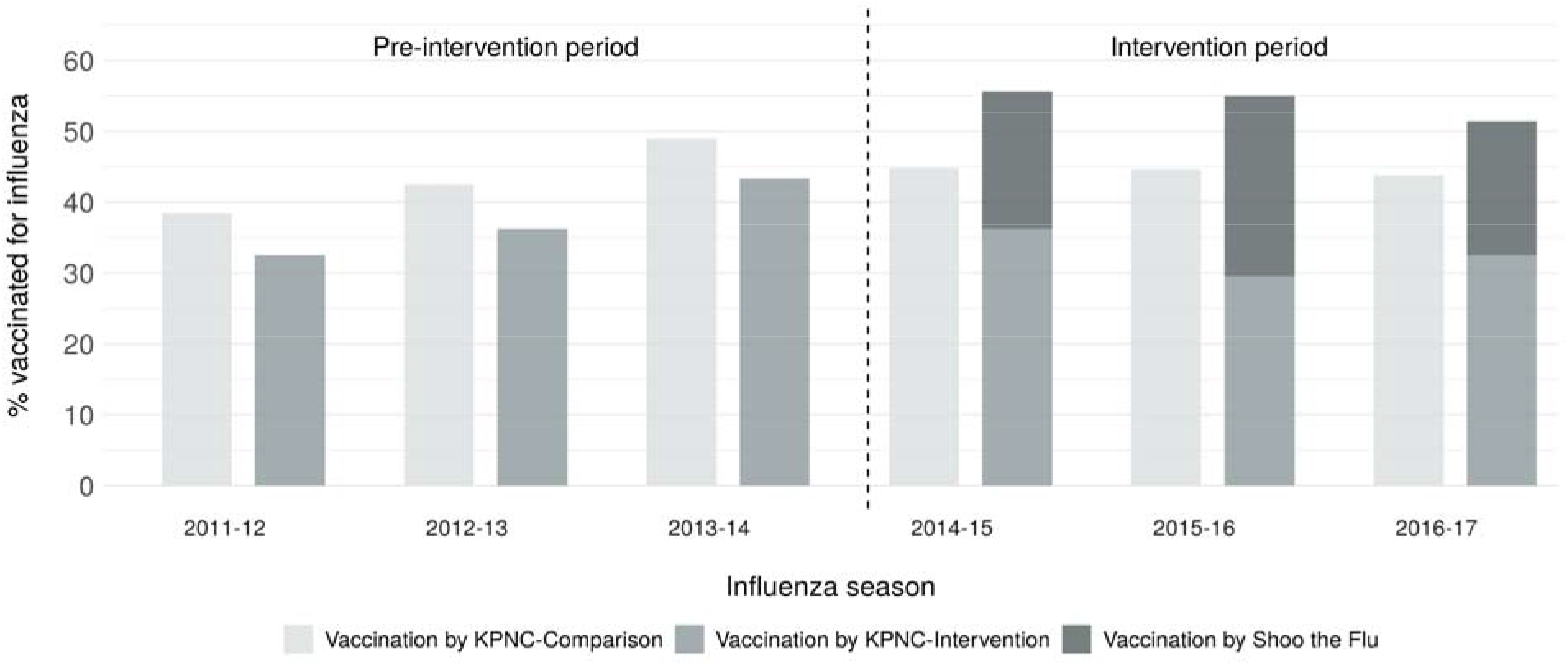
**Percentage of elementary school aged study population vaccinated for influenza by Kaiser Permanente Northern California or the Shoo the Flu intervention** “Vaccination by KPNC – Comparison” includes influenza vaccinations delivered by KPNC in the comparison site. “Vaccination by KPNC – Intervention” includes influenza vaccinations delivered by KPNC in the intervention site. “Vaccination by Shoo the Flu” includes influenza vaccinations delivered by the Shoo the Flu intervention in the intervention site. The percentage vaccinated for influenza does not include vaccinations that were not delivered by KPNC or Shoo the Flu. The denominator is KPNC patients aged 5-12 years.

### Cumulative incidence of influenza-related outcomes

The cumulative incidence of MAARI ranged from 0.14 to 0.58 per season by age and was highest among children 0-4 years (Supplement Figure 5). Very few individuals were tested for influenza (<1% of individuals per season), and the proportion who tested positive varied substantially by age, season, and site. The incidence of influenza hospitalization was low in all age groups, with the highest incidence among adults ≥65 years (range: 0.053 to 0.097 per season). The incidence of filled Oseltamivir prescriptions ranged from 0.0007 to 0.0134 per season.

### Associations among elementary school aged children

Overall, the intervention was not associated with MAARI in elementary school aged children except for in 2015-16, when it was associated with lower MAARI when accounting for pre-season differences between sites (Figure 2, Supplement Figure 6, Supplement Table 5). Among elementary school aged children, in 2016-17 the unadjusted pre-intervention DID in the cumulative incidence of filled Oseltamivir prescriptions per 1,000 was −3.5 (95% CI −5.5, −1.5) in 2015-16 and −4.0 (95% CI −6.5, −1.6) in 2016-17; there was no association in 2014-15. These associations were attenuated towards the null in the analysis accounting for pre-season differences. There was no association with influenza hospitalization elementary school aged children.

**Figure 2.**
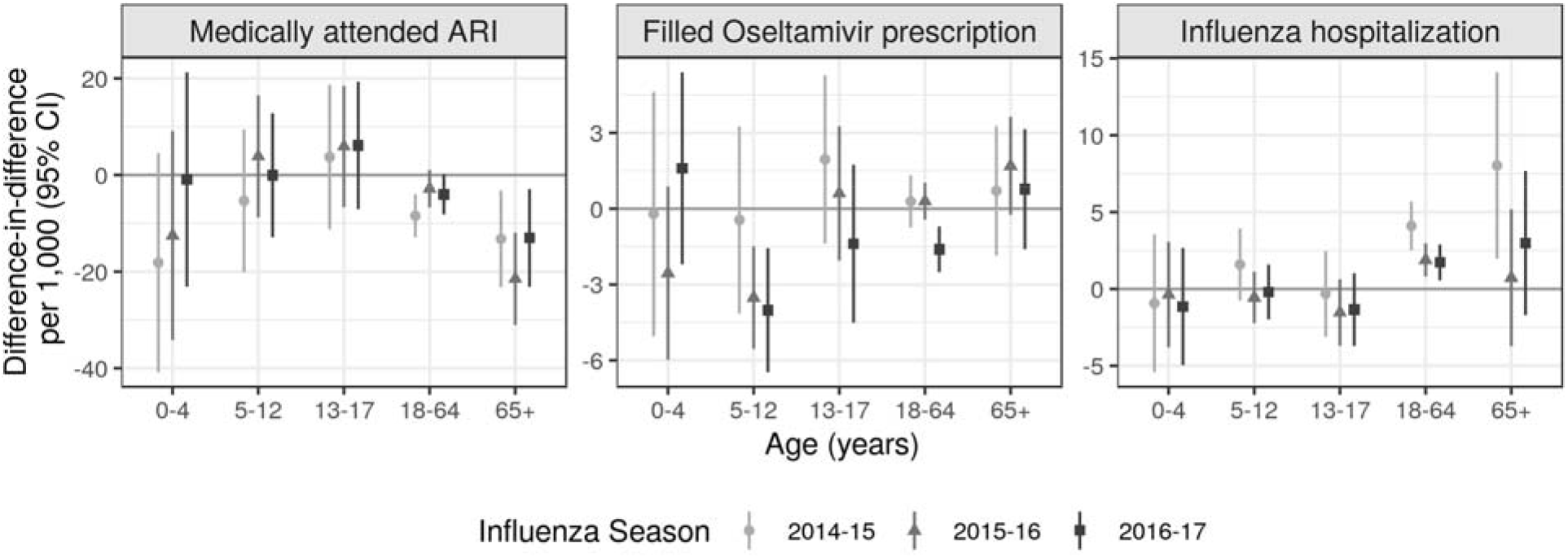
Difference-in-differences accounting for pre-intervention differences. Difference-in-difference in cumulative incidence of each outcome during each influenza season comparing the difference in mean outcome in each district in an intervention year compared to the three pre-intervention years (2011-2013). Difference-in-difference parameters remove any time-invariant differences between groups (measured or unmeasured). Parameters were estimated using a generalized linear model without covariate adjustment due to data sparsity. Standard errors accounted for clustering at the household level. Estimates in children 5-12 years measure total effects and estimates in other age groups measure indirect effects. Analyses were restricted to influenza season defined based the percentage of medical visits for influenza-like illness in California as reported by the California Department of Public Health. Influenza season started when there were at least 2 consecutive weeks in which the percentage of medical visits for influenza-like illness exceeded 2%, and the season ended when there were at least two consecutive weeks in which the percentage was less than or equal to 2%.

### Associations among non-elementary school aged individuals

DIDs indicated no association between the intervention and laboratory-confirmed influenza in most program years and age groups among non-elementary school aged individuals (Supplement Table 5). In intervention vs. comparison sites, MAARI per 1,000 was lower among adults 65 years or older when accounting for pre-intervention differences (2014-15: −13.2, 95% CI −23.2, −3.2; 2015-16: −21.5, 95% CI −31.1, −11.9; 2016-17: −13.0, 95% CI −23.2, −2.9) (Figure 2). When accounting for pre-season differences, DIDs indicated slightly stronger protective associations with MAARI for most age groups and seasons (Supplement Figure 6). DIDs for filled Oseltamivir prescriptions per 1,000 were lower among adults 18-64 years in 2016-17 (−1.6, 95% CI −2.5, −0.7); results were similar when accounting for pre-season differences. DIDs indicated a higher incidence of influenza hospitalization among adults 18 years or older in 2014-15 and adults 18-64 years in 2015-16. Pre-season DIDs displayed a different pattern for adults: associations were null for adults 18 years or older except for adults 65 years or older in 2015-16, when the association was protective.

For all ages, the results of adjusted models were similar to those of unadjusted models (Supplement Table 6). Results stratified by vaccination status were similar overall to the primary analysis (Supplement Tables 7-8).

### Negative control analyses

Analyses using non-influenza outcomes (medically attended diarrhea and medically attended gastrointestinal illness) produced primarily null associations (Supplement Table 9). Analyses restricted to weeks outside of influenza season produced almost exclusively null results (Supplement Table 10).

### Sensitivity analyses using alternative influenza season definitions

Results were similar when using alternative influenza season definitions (Supplement Figure 7). For outcomes with sufficient data to restrict to the peak week of influenza, results suggested no association with the intervention.

## Discussion

In this three-year evaluation of a city-wide SLIV intervention, among elementary school aged children, SLIV was associated with lower Oseltamivir prescriptions but not with MAARI, laboratory-confirmed influenza, or influenza hospitalization. We found some evidence of indirect effects in non-elementary school aged individuals: SLIV was associated with lower MAARI among adults 65 years or older and lower Oseltamivir prescriptions among pre-school children in 2015-16.

Some prior observational studies of SLIV programs have also reported indirect effects of SLIV on MAARI and related outcomes [14,18–20], while others have reported non-significant or null results [7,8,13,16,17]. These studies did not use analytic methods to account for differences between intervention and comparison sites prior to intervention or outside of influenza season. The one prior study that did so found no association between SLIV and hospitalization among the elderly [16]. The present study included three years of pre-intervention data and used a DID approach to adjust for pre-intervention differences; thus, our findings likely have higher internal validity than prior observational SLIV evaluations.

Vaccine effectiveness varied substantially during the study period. In 2014-15 all vaccine formulations had low effectiveness due to a poor strain match [24]. In 2015-16, the LAIV had poor vaccine effectiveness, but the IIV was moderately effective [26]. In 2016-17, only IIV was available and it was moderately effective [35]. These differences and varying vaccine coverage by the SLIV intervention each year [21] contribute to heterogeneous estimates across seasons.

SLIV may increase vaccination coverage among children who would not otherwise be vaccinated and/or shift vaccination location from doctor’s offices to schools. If SLIV merely shifts vaccination location, it would not be expected to reduce influenza. In this study, SLIV appeared to both increase vaccination and shift vaccination location among elementary school aged children. The proportion of 5-12 year-olds vaccinated by their medical provider was lower in the intervention site than the comparison site, but the proportion vaccinated by KPNC or SLIV combined was higher. This finding is consistent with our prior evaluation of Shoo the Flu [21] and evaluations of other SLIV interventions that also increased vaccination coverage [4–13].

Our finding that SLIV was associated with higher influenza hospitalization in adults in some years was unexpected. This finding contrasts with the prior Shoo the Flu evaluation, which found that SLIV was associated with lower influenza hospitalization in non-elementary age groups [21]. Other prior SLIV evaluations have had conflicting findings with regard to indirect effects on hospitalization [7,8,13,16,17]. One cluster-randomized trial reported a higher hospitalization rate in SLIV participants, but when stratifying by child vaccination status, the finding was not significant [4]. In a post-hoc analysis accounting for pre-season differences between sites, which may better account for time-dependent confounding than our primary analysis, associations with hospitalization were null or protective.

This study is subject to several limitations. First, its observational design may be subject to unmeasured confounding. In a small number of negative control outcome and time period analyses, unexpected associations suggest that unmeasured confounding occurred. The DID analysis controlled for time-invariant confounding [28], but unmeasured time-varying confounding may have occurred. Common confounders of influenza vaccine studies are age, calendar time, and health status [36]. Our analysis stratified by age and season, but baseline health status was not available. Second, differences in sociodemographic characteristics between the study population and the general and student populations in the study sites limit the generalizability of our findings. Third, many outcomes were rare, precluding formal analyses for some outcomes and limiting statistical power, which may have contributed to null findings. Finally, we did not have complete vaccination information about each individual. It was not possible to link data from the California Immunization Registry with KPNC member data. This limitation underscores the need for more robust vaccine registries [37]. Notably, the limitations of this study apply to most prior SLIV evaluations, which primarily have been observational and leveraged existing data. We expect that among individuals not targeted by the SLIV intervention, influenza vaccinations outside KPNC were infrequent, so this limitation is unlikely to have meaningfully impacted study findings.

Overall, our findings bolster those of our prior evaluation of a city-wide SLIV intervention. This study supports our prior finding that SLIV both increased vaccination coverage and shifted vaccination location and provides additional evidence that SLIV was associated with lower Oseltamivir prescriptions in school aged children and lower MAARI in older adults.

## Data Availability

The data from this study is not available due to human subjects protections.

## Funding

This work was supported by a grant from the Flu Lab (https://theflulab.org/) to the University of California, Berkeley (Grant number: 20142281; PI: AR).

## Acknowledgements

The authors would like to acknowledge Kristin Goddard, Karen Nunley, and Berwick Chan at Kaiser Permanente Northern California and Kate Holbrook at Alameda County Public Health Department for their contributions to data collection.

NPK reports research support from Protein Science (Sanofi Pasteur) and Sanofi Pasteur for unrelated influenza vaccine studies and from GlaxoSmithKline, Merck & Co, and Pfizer for unrelated studies. JBC, BFA, KM, CJK, AN, NNP, SD, AS, AEH, AR, JMC report no conflicts.

