## Supplement for "City-wide school-located influenza vaccination: a retrospective cohort study"

**Supplement 1. Additional details about the Shoo the Flu intervention**

The intervention aimed to increase influenza vaccination coverage of elementary schoolchildren with the goal of reducing influenza transmission in elementary schoolchildren and community-wide through indirect effects. Children were eligible for vaccination regardless of their insurance status.

In 2014-15 and 2015-16, the intervention implemented a mass media campaign in Oakland, California that included advertisements in subways, bus shelters, billboards, newspapers, and digital media. Though these promotion efforts were restricted to the Oakland area, some residents of other nearby areas may have been exposed to intervention promotion media.

**Supplement 2. Additional study design details**

Selection of comparison district

We conducted a retrospective cohort study of KPNC health plan members who lived in the catchment areas of the intervention district and a comparison district. To identify the comparison site, we considered school districts in the San Francisco Bay Area with five or more elementary schools located at least five miles from the intervention district. We used a genetic multivariate matching algorithm [28] to pair-match public elementary schools in the intervention district and each candidate comparison district using pre-intervention school-level characteristics: mean enrollment, class size, parental education, academic performance index scores, California standardized test scores, and the school-level percentage of English language learners and students receiving free lunch at school. We selected West Contra Costa Unified School District (WCCUSD) as the comparison site because it had the smallest average generalized Mahalanobis distance between paired schools [28].

Study size

Since our analysis was limited to the KPNC study population, to assess statistical power, we estimated minimum detectable relative risks assuming a two-sided Type I error of 0.05 (Hayes and Bennett, 1999). We calculated the intraclass correlation coefficient for the household-level clustering for each outcome and accounted for these ICCs in the power calculation. There was at least 80% statistical power to detect the following minimum relative risks: MAARI (0.93), positive influenza test (0.93), influenza hospitalization (0.80), and Oseltamivir prescription (0.63).

Hayes RJ, Bennett S. Simple sample size calculation for cluster-randomized trials. Int J Epidemiol **1999**; 28:319–326.

**Supplement 3. Deviations from pre-analysis plan**

Although Oseltamivir prescriptions (regardless of whether they were filled) was also a pre-specified outcome, we only present results for filled Oseltamivir prescriptions because each influenza season, fewer than 5 patients who received a prescription did not fill it.

We pre-specified laboratory-confirmed respiratory syncytial virus infection as a negative control outcome, but it did not meet the criterion of at least 10 outcome events per variable.

**Supplement 4. Additional analysis details**

Our primary analysis pooled across individual influenza vaccination status. Since individuals who were vaccinated for influenza at KPNC were unlikely to have participated in the SLIV intervention, to further isolate the effect of the intervention we performed a pre-specified secondary analysis that stratified by whether individuals were vaccinated for influenza at KPNC. Individual-level vaccination data for vaccinations from sources other than KPNC, including the SLIV intervention, was not available. Individuals not vaccinated for influenza at KPNC may have received the influenza vaccine at school or another source or may have been unvaccinated. As such, estimates among the individuals not vaccinated at KPNC capture both direct effects of SLIV and indirect effects among the unvaccinated. Estimates among individuals vaccinated at KPNC capture potential indirect effects of SLIV on non-participants in the SLIV intervention.

Analyses assumed all missing covariate data were missing completely at random. We also used targeted maximum likelihood estimation results (Laan & Rose, 2011) were similar, so we report generalized linear models results.

van der Laan, M. J. & Rose, S. *Targeted Learning: Causal Inference for Observational and Experimental Data*. (Springer, 2011).

**Supplement Figure 1. Definition of total effects and indirect effects in this study**

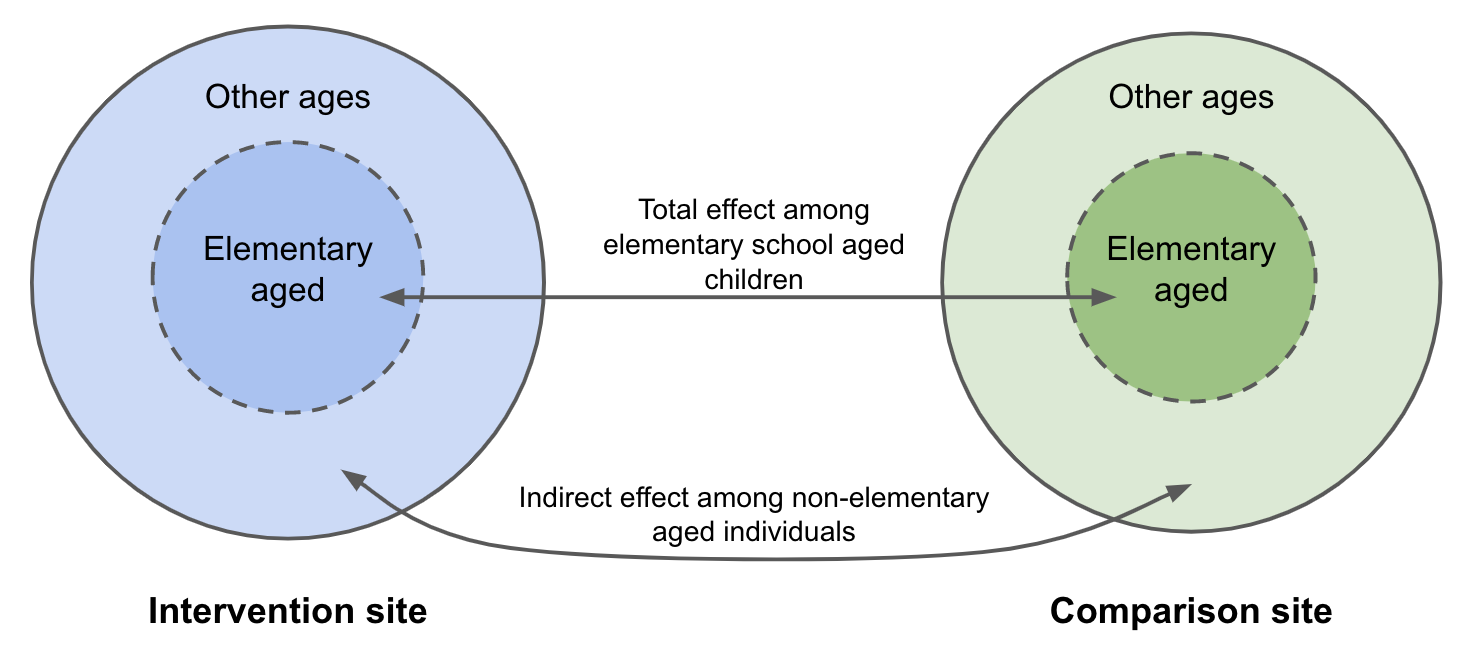

The total effect of the SLIV intervention compares outcomes between elementary school aged children in the intervention vs. comparison site; it combines the direct effect among children who attended intervention schools and the indirect effect among children who did not attend intervention schools. Because it was not possible to link school information to Kaiser patient data, we were not able to decompose total effects into direct and indirect effects among elementary school aged children. The indirect effect among non-elementary aged individuals compares outcomes between individuals in the intervention site (who were not eligible for the SLIV intervention) to those of non-elementary aged individuals in the comparison site.

**Supplement Figure 2. Monthly cumulative incidence of each outcome by site**

**A. Medically attended acute respiratory infection**

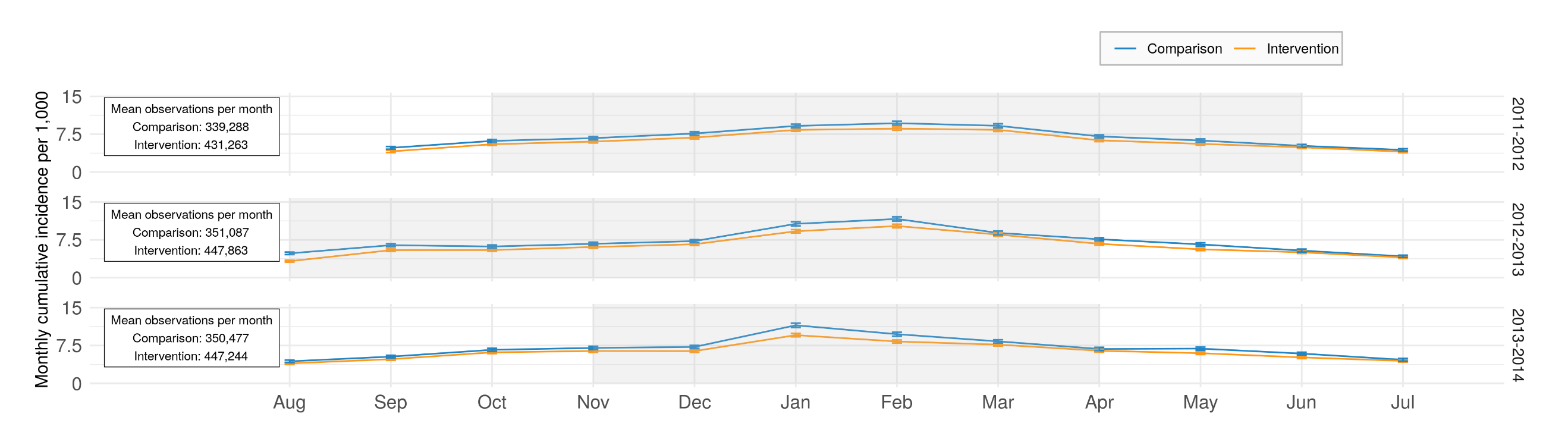

**B. Laboratory-confirmed influenza infection among tested individuals**

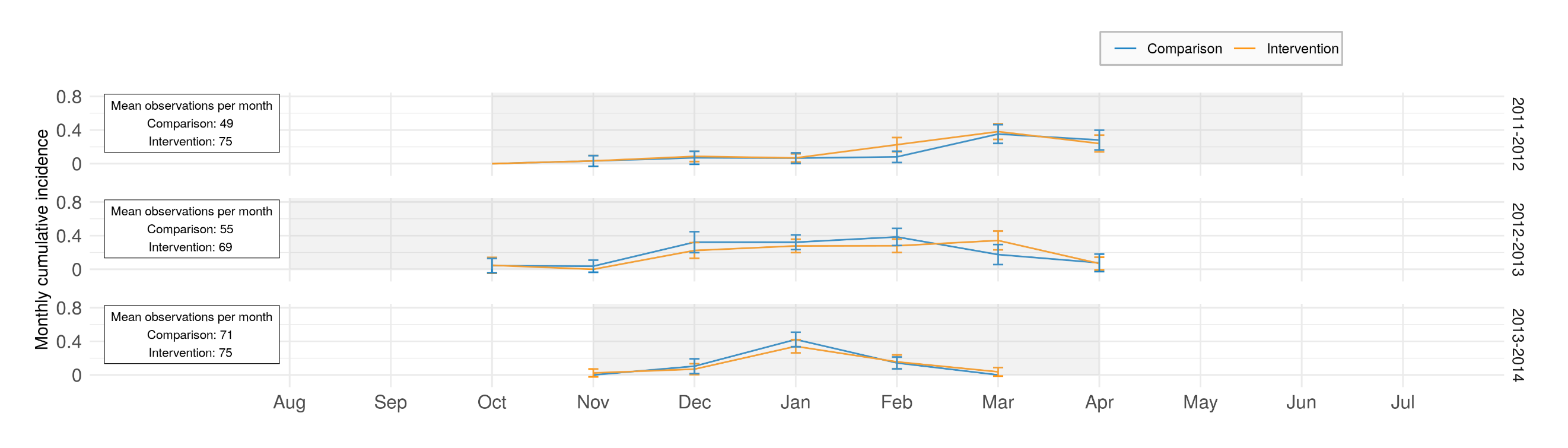

**C. Influenza hospitalization**

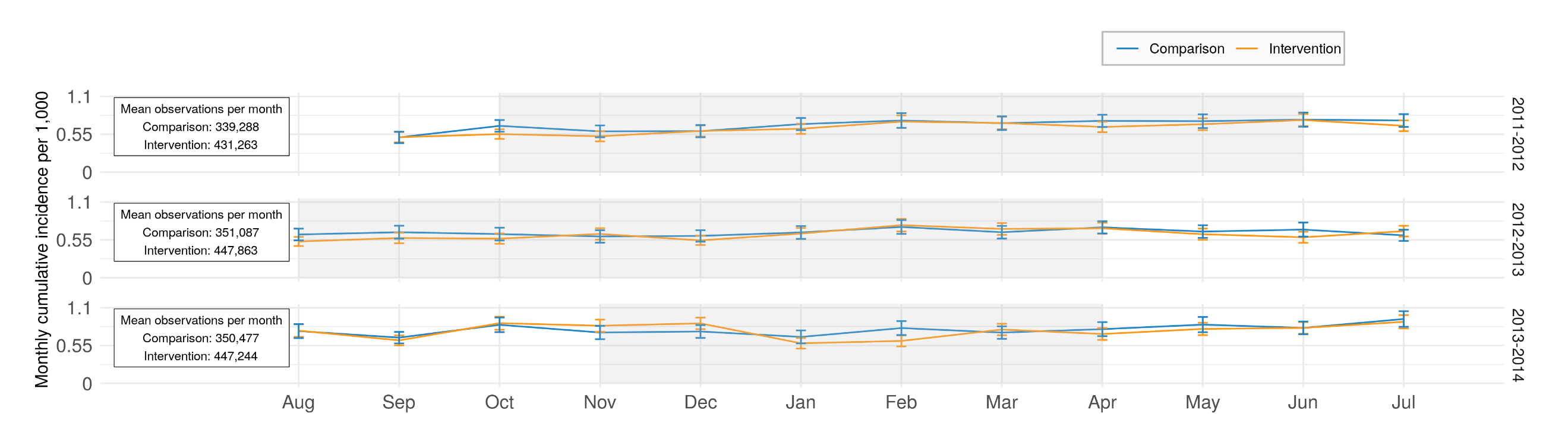

**D. Filled Oseltamivir prescriptions**

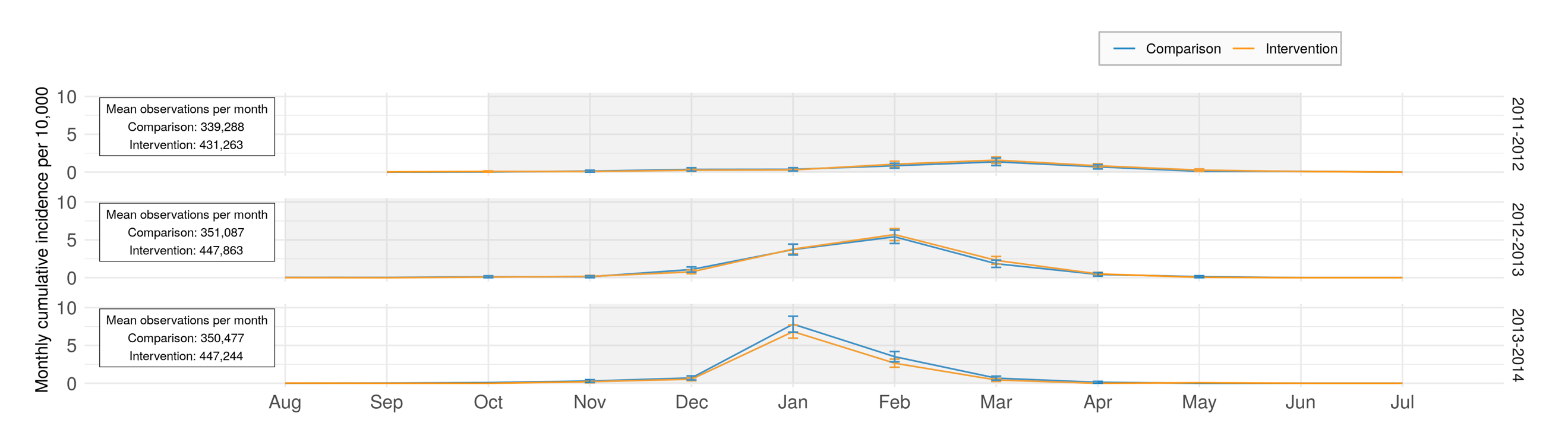

**E. Medically attended diarrhea (negative control outcome)**

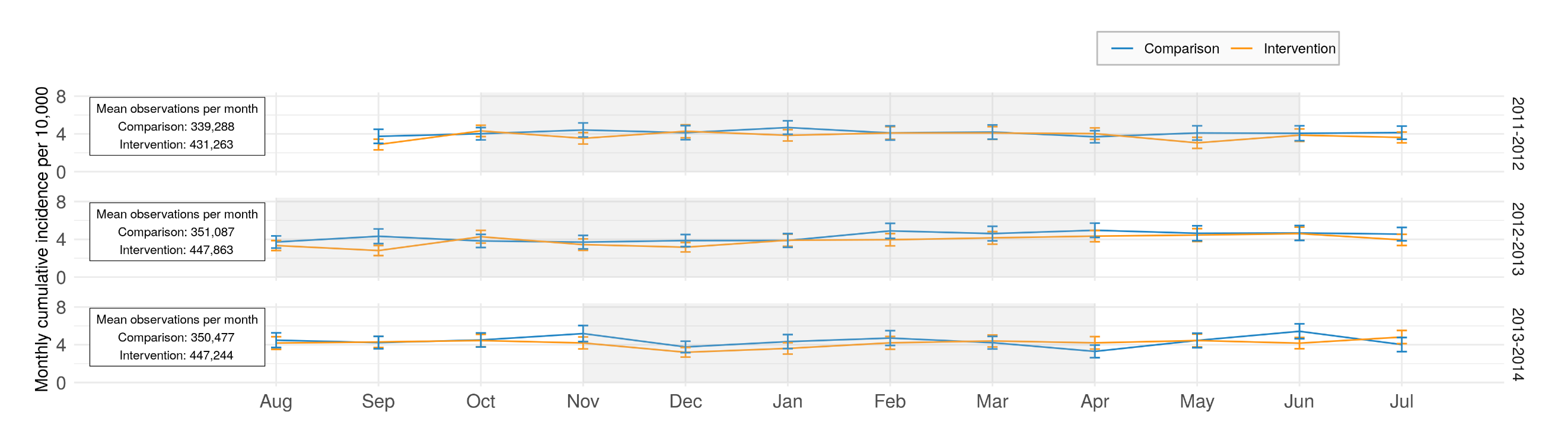

**F. Medically attended gastrointestinal illness (negative control outcome)**
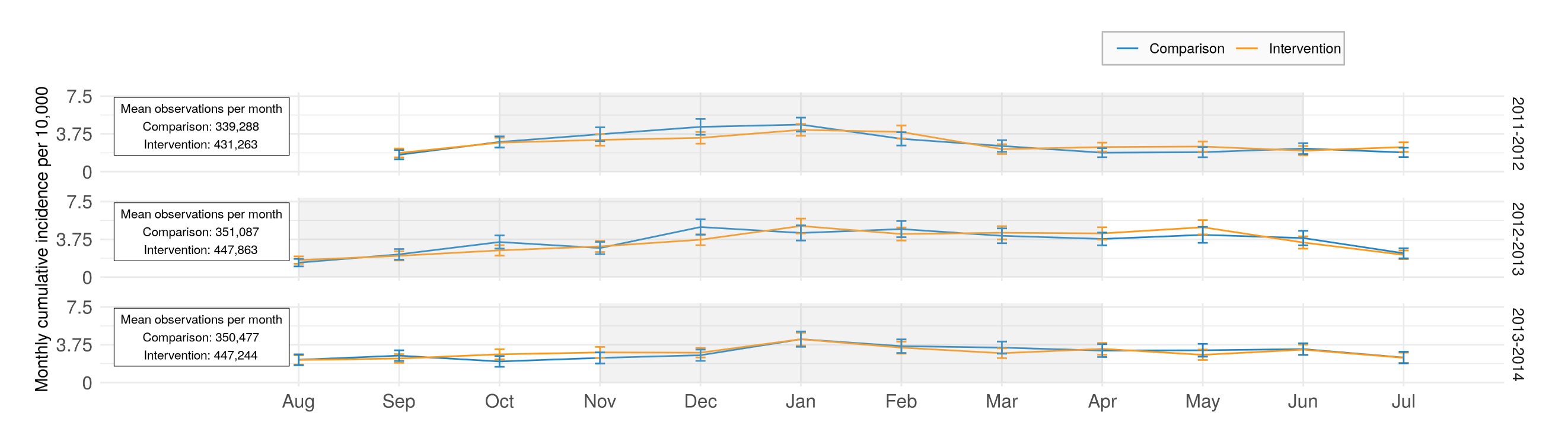

**Supplement Figure 3. Influenza vaccination at KPNC by age group and influenza season**

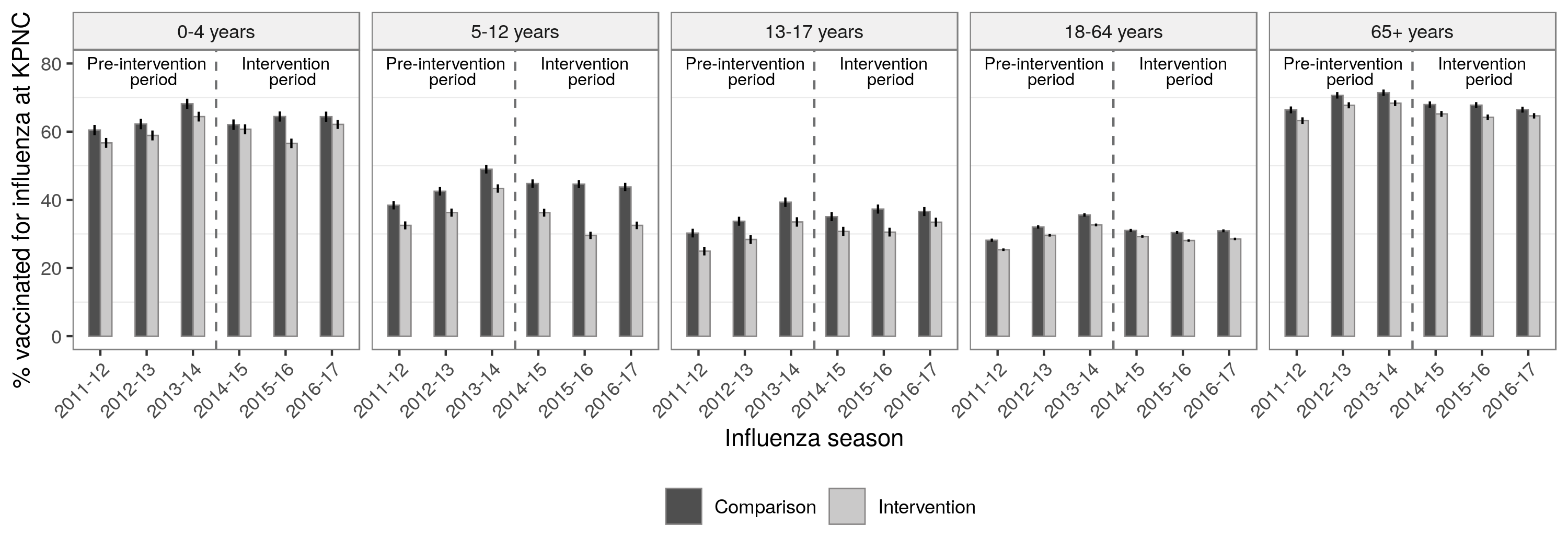

Includes all influenza vaccinations at KPNC between September 1 and August 31 each year.

**Supplement Figure 4. Influenza vaccination at KPNC by vaccine type, age group, and influenza season**

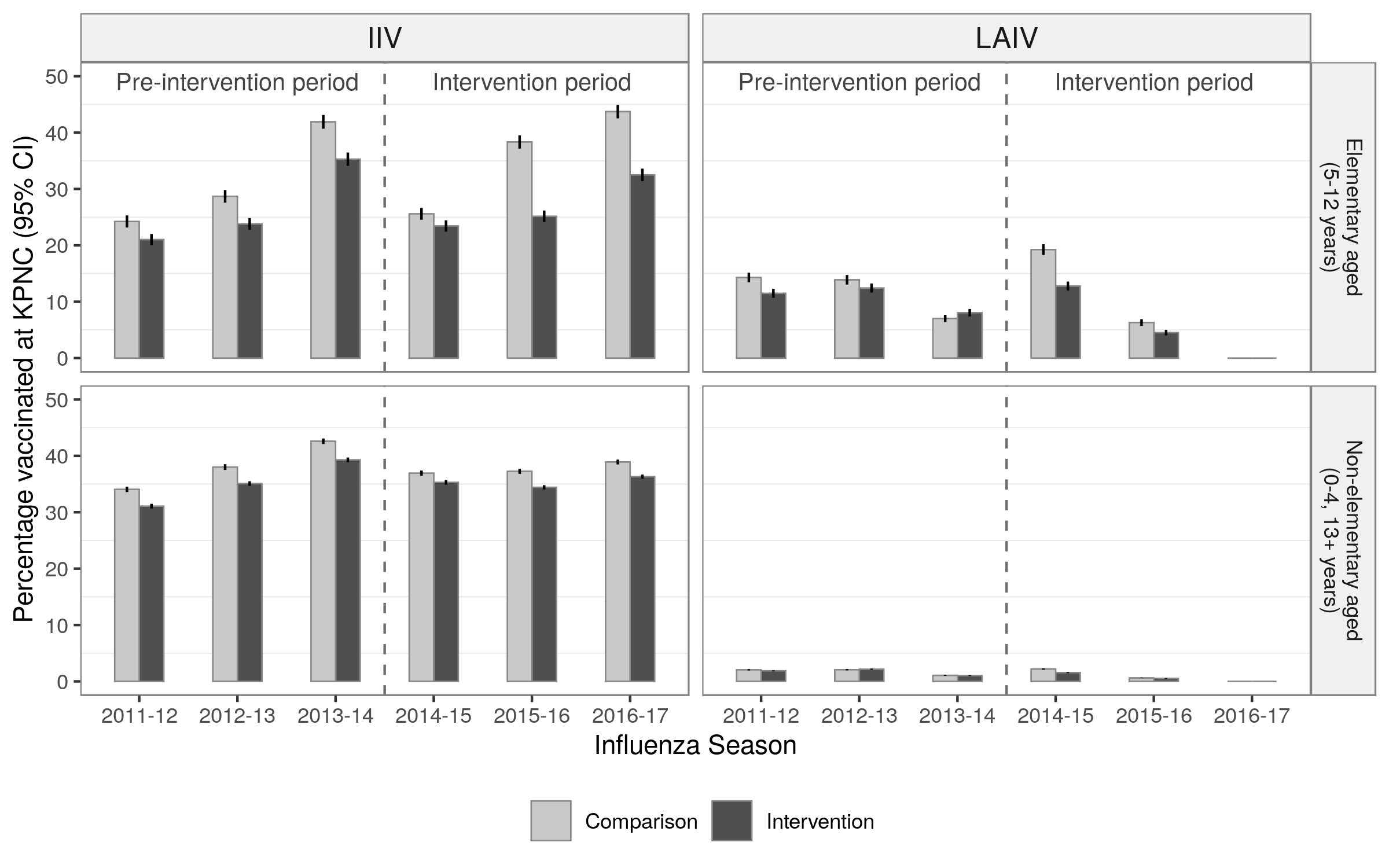

Includes all influenza vaccinations at KPNC between September 1 and August 31 each year.

**Supplement Figure 5. Cumulative incidence of each outcome during influenza season by age category and season**

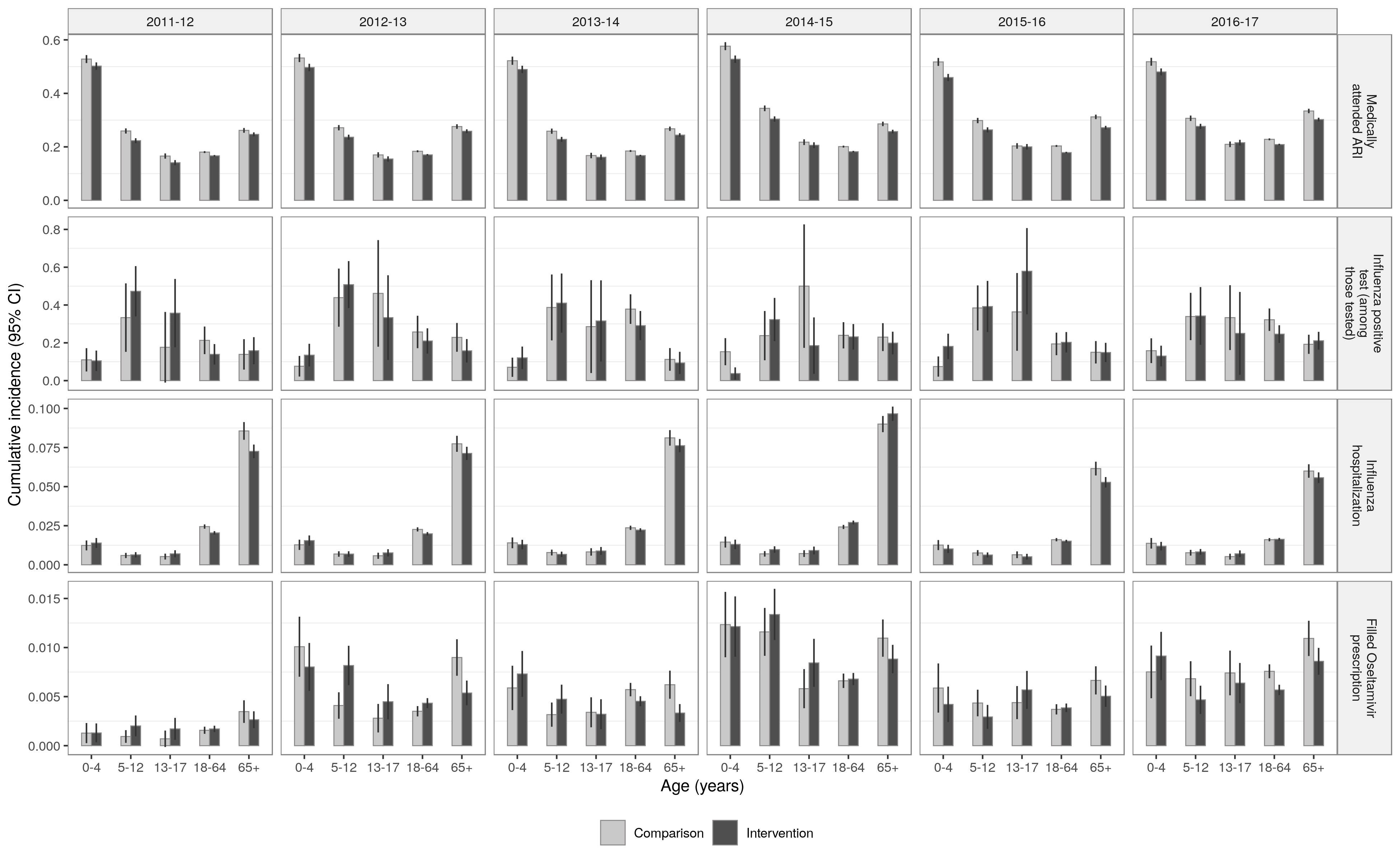

Influenza seasons 2011-12, 2012-13, and 2013-14 were prior to the intervention, and seasons 2014-15, 2015-16, 2016-17 were during the intervention period. Vertical lines in the center of each bar indicate the 95% confidence intervals calculated with robust standard errors to account for clustering at the household level. The standard errors for influenza positive tests did not adjust for clustering because there were very few households with multiple positive individuals. Cumulative incidence was estimated within influenza season defined based the percentage of medical visits for influenza-like illness in California as reported by the California Department of Public Health. Influenza season started when there were at least 2 consecutive weeks in which the percentage of medical visits for influenza-like illness exceeded 2%, and the season ended when there were at least two consecutive weeks in which the percentage was less than or equal to 2%.

**Supplement Figure 6. Difference-in-differences accounting for pre-season differences**

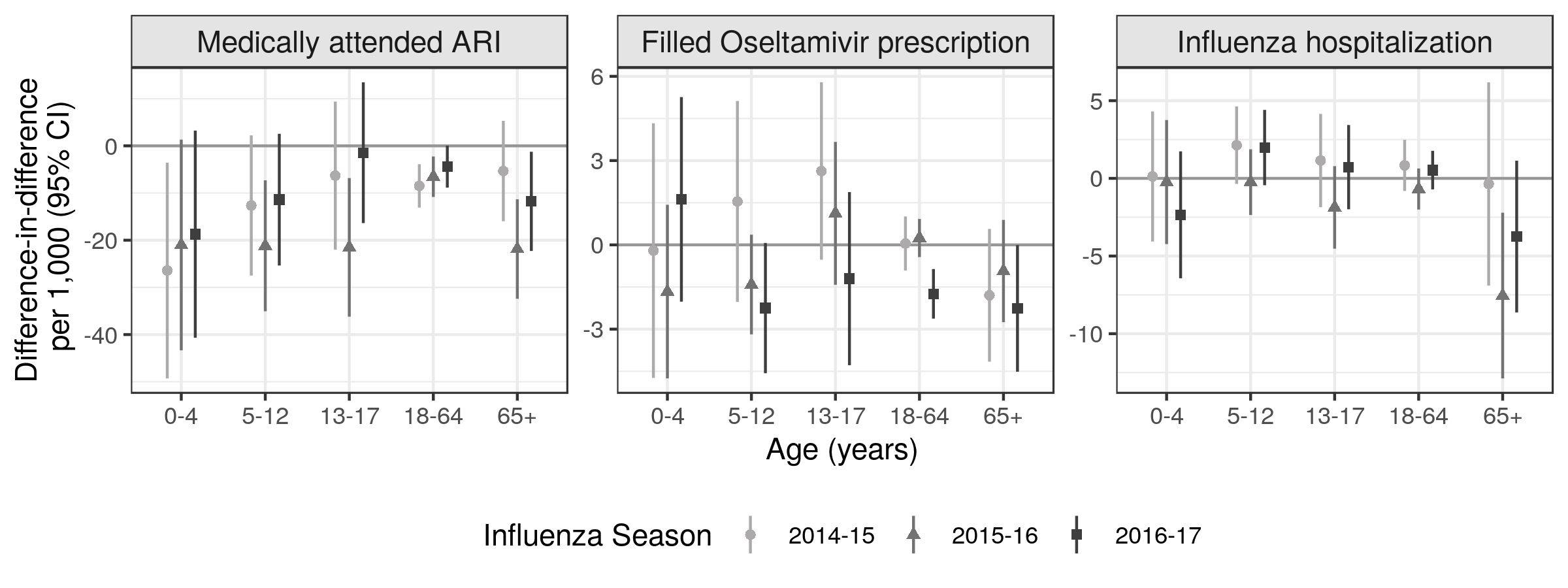

Difference-in-difference comparing the difference in mean outcome in each district in an intervention year compared to the three months prior to each season (May through September). Parameters were estimated using a generalized linear model without covariate adjustment due to data sparsity. Standard errors accounted for clustering at the household level. Estimates in children 5-12 years measure total effects, and estimates in other age groups measure indirect effects. Analyses were restricted to influenza season defined based the percentage of medical visits for influenza-like illness in California as reported by the California Department of Public Health. Influenza season started when there were at least 2 consecutive weeks in which the percentage of medical visits for influenza-like illness exceeded 2%, and the season ended when there were at least two consecutive weeks in which the percentage was less than or equal to 2%.

**Supplement Figure 7. Sensitivity analyses with alternative influenza season definitions**

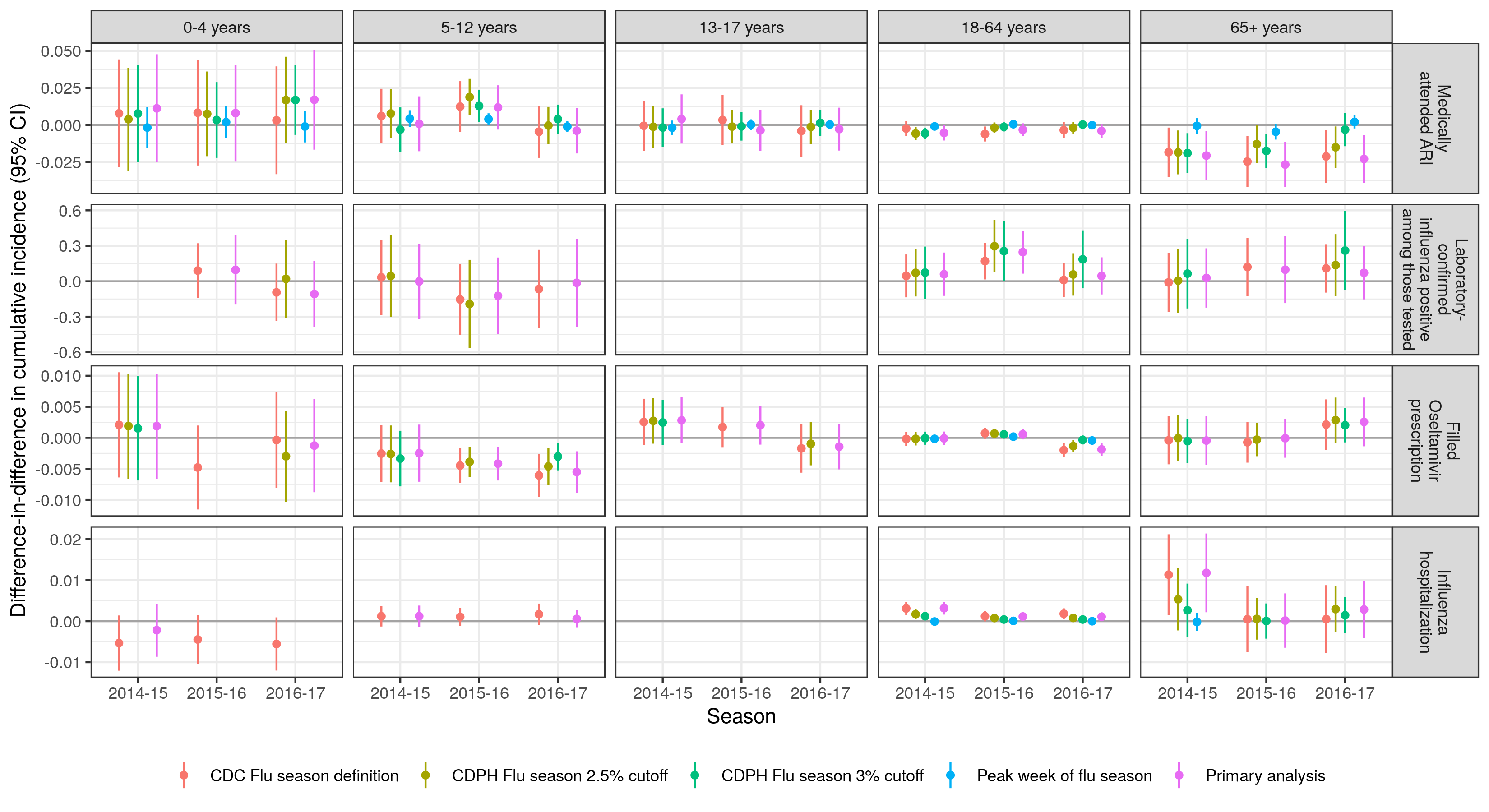

Analyses were restricted to influenza season defined based the percentage of medical visits for influenza-like illness in California as reported by the California Department of Public Health. Influenza season started when there were at least 2 consecutive weeks in which the percentage of medical visits for influenza-like illness exceeded 2%, and the season ended when there were at least two consecutive weeks in which the percentage was less than or equal to 2%. Difference-in-difference estimates compare the difference in mean outcome in each district in an intervention year compared to the three pre-intervention years (2011-2013). Difference-in-difference parameters remove any time-invariant differences between groups (measured or unmeasured). Parameters were estimated using a generalized linear model without covariate adjustment due to data sparsity. Standard errors accounted for clustering at the household level. Estimates are not shown for some outcomes and ages due to sparse data.

**Supplement Table 1. International Classification of Diseases, Clinical Modification, Version 9 and 10 (ICD 9/10) codes used to define medically attended acute respiratory illness visits**

| **CODE GROUP** | **ICD 9 CODE** | **CONDITION** |
| --- | --- | --- |
| Viral and chlamydial infection in conditions classified elsewhere and of unspecified site | 79.99 | Unspecified viral infection |
| Diseases of ear and mastoid process | 382.0* | Acute suppurative otitis media |
| Diseases of ear and mastoid process | 382.4* | Unspecified suppurative otitis media |
| Diseases of ear and mastoid process | 382.9* | Unspecified otitis media |
| Acute respiratory infections | 460.* | Acute nasopharyngitis (common cold) |
| Acute respiratory infections | 461.* | Acute sinusitis |
| Acute respiratory infections | 464.* | Acute laryngitis and tracheitis |
| Acute respiratory infections | 465.* | Acute upper respiratory infections of multiple or unspecified sites |
| Acute respiratory infections | 466.* | Acute bronchitis and bronchiolitis |
| Pneumonia and influenza | 480.* | Viral pneumonia |
| Pneumonia and influenza | 483.* | Pneumonia due to other specified organism |
| Pneumonia and influenza | 484.* | Pneumonia in infectious diseases classified elsewhere |
| Pneumonia and influenza | 485.* | Bronchopneumonia, organism unspecified |
| Pneumonia and influenza | 486.* | Pneumonia, organism unspecified |
| Pneumonia and influenza | 487.* | Influenza |
| Pneumonia and influenza | 488.* | Influenza due to identified avian influenza virus |
| Chronic obstructive pulmonary disease and allied conditions | 490.* | Bronchitis, not specified as acute or chronic |
| Chronic obstructive pulmonary disease and allied conditions | 491.* | Chronic Bronchitis |
| Chronic obstructive pulmonary disease and allied conditions | 493.* | Asthma |
| General symptoms | 780.60 | Fever, unspecified |
| Symptoms involving respiratory system and other chest symptoms | 786.2* | Cough |
| Dyspnea and respiratory abnormalities | 786.07 | Non-specific wheezing; excludes asthma |
| **Code Group** | **ICD10 code** | **Condition** |
| Acute respiratory infections | J00 | Acute nasopharyngitis [common cold] |
| Acute respiratory infections | J01.* | Acute sinusitis |
| Acute respiratory infections | J02.* | Acute pharyngitis |
| Acute respiratory infections | J03.* | Acute tonsillitis |
| Acute respiratory infections | J04.* | Acute laryngitis and tracheitis |
| Acute respiratory infections | J05.* | Acute obstructive laryngitis [croup] and epiglottitis |
| Acute respiratory infections | J06.* | Acute upper respiratory infections of multiple and unspecified sites |
| Acute respiratory infections | J20.* | Acute bronchitis |
| Acute respiratory infections | J21.* | Acute bronchiolitis |
| Chronic obstructive pulmonary disease and allied conditions | J40.* | Bronchitis, not specified as acute or chronic |
| Chronic obstructive pulmonary disease and allied conditions | J41.* | Simple and mucopurulent chronic bronchitis |
| Chronic obstructive pulmonary disease and allied conditions | J42.* | Unspecified chronic bronchitis |
| Chronic obstructive pulmonary disease and allied conditions | J44.* | Other chronic obstructive pulmonary disease |
| Chronic obstructive pulmonary disease and allied conditions | J45.* | Asthma |
| Diseases of ear and mastoid process | H66.0* | Acute suppurative otitis media |
| Diseases of ear and mastoid process | H66.4* | Suppurative otitis media, unspecified |
| Diseases of ear and mastoid process | H66.9 | Otitis media, unspecified |
| Diseases of ear and mastoid process | H67.* | Otitis media in diseases classified elsewhere |
| Dyspnea and respiratory abnormalities | R06.2 | Wheezing |
| General symptoms | R50.9 | Fever, unspecified |
| General symptoms | R68.83 | Chills (without fever) |
| Pneumonia and influenza | A37.01 | Whooping cough due to Bordetella pertussis with pneumonia |
| Pneumonia and influenza | A37.11 | Whooping cough due to Bordetella parapertussis with pneumonia |
| Pneumonia and influenza | A37.81 | Whooping cough due to other Bordetella species with pneumonia |
| Pneumonia and influenza | A37.91 | Whooping cough, unspecified species with pneumonia |
| Pneumonia and influenza | B25.0 | Cytomegaloviral pneumonitis |
| Pneumonia and influenza | J09.* | Influenza due to certain identified influenza viruses |
| Pneumonia and influenza | J10.* | Influenza due to other identified influenza virus |
| Pneumonia and influenza | J11.* | Influenza due to unidentified influenza virus |
| Pneumonia and influenza | J12.* | Viral pneumonia, not elsewhere classified |
| Pneumonia and influenza | J13.* | Pneumonia due to Streptococcus pneumoniae |
| Pneumonia and influenza | J14.* | Pneumonia due to Hemophilus influenzae |
| Pneumonia and influenza | J15.* | Bacterial pneumonia, not elsewhere classified |
| Pneumonia and influenza | J16.* | Pneumonia due to other infectious organisms, not elsewhere classified |
| Pneumonia and influenza | J17.* | Pneumonia in diseases classified elsewhere |
| Pneumonia and influenza | J18.* | Pneumonia, unspecified organism |
| Symptoms involving respiratory system and other chest symptoms | R05 | Cough |

**Supplement Table 2. Number of patients and person-weeks by year and season**

|  | Number of patients | Person-weeks | |
| --- | --- | --- | --- |
|  |  | Entire follow-up period | Influenza season |
| 2011-12 | 175,628 | 9,436,202 | 6,801,780 |
| 2012-13 | 226,875 | 9,623,350 | 5,861,011 |
| 2013-14 | 229,312 | 9,748,880 | 3,991,309 |
| 2014-15 | 240,844 | 10,503,001 | 5,981,703 |
| 2015-16 | 254,826 | 11,138,348 | 3,069,633 |
| 2016-17 | 269,266 | 11,500,570 | 3,640,907 |

The entire follow-up period was from September 1 to August 31 of the following calendar year. Influenza season was defined based the percentage of medical visits for influenza-like illness in California as reported by the California Department of Public Health. Influenza season started when there were at least 2 consecutive weeks in which the percentage of medical visits for influenza-like illness exceeded 2%, and the season ended when there were at least two consecutive weeks in which the percentage was less than or equal to 2%.

**Supplement Table 3. Pre-intervention characteristics of study population, general population, and school district population**

|  | Study Population ^a^ | | General Population ^b^ | | School District Population ^c^ | |
| --- | --- | --- | --- | --- | --- | --- |
|  | Intervention (%) | Comparison (%) | Intervention (%) | Comparison (%) | Intervention (%) | Comparison (%) |
| Age (years) |  |  |  |  |  |  |
| Under 5 | 5 | 6 | 7 | 7 | -- | -- |
| 5 to 14 | 11 | 14 | 11 | 13 | -- | -- |
| 15 to 17 | 3 | 4 | 3 | 4 | -- | -- |
| 18 to 64 | 66 | 62 | 68 | 65 | -- | -- |
| 65 + | 15 | 15 | 11 | 12 | -- | -- |
| Race |  |  |  |  |  |  |
| Asian / Pacific Islander | 15 | 20 | 17 | 19 | 15 | 16 |
| Black / African American | 28 | 20 | 28 | 19 | 18 | 27 |
| Native American | 0 | 0 | 1 | 1 | 0 | 0 |
| Multiracial | 1 | 1 | 6 | 6 | 1 | 3 |
| White | 35 | 30 | 35 | 37 | 11 | 13 |
| Other ^d^ | -- | -- | 14 | 18 | -- | -- |
| Unknown or not reported | 20 | 29 | 1 | 1 | 0 | 1 |
| Hispanic ethnicity / race ^e^ | 31 | 43 | 26 | 33 | 54 | 39 |
| MediCAL enrollee | 4 | 7 | 2 | 1 | -- | -- |
| Primary language spoken is not English | 12 | 14 | 40 | 47 | 43 | 38 |

^a^ KPNC patient population between September 1, 2013 and August 31, 2014

^b^ U.S. 2010 Census data for zip codes overlapping with the study site

^c^ California Department of Education data from 2013

^d^ This category was not included in the Kaiser or California Department of Education datasets.

^e^ KPNC data and the Census consider Hispanic to be an ethnicity, while the California Department of Education data considered it a race. Thus, race category percentages sum to 100% excluding Hispanic for the Study Population and the General Population, and they sum to 100% including Hispanic for the School District Population.

**Supplement Table 4. Percentage of elementary school aged patients vaccinated for influenza at KPNC during each year of the study**

|  | **Intervention** | | **Comparison** | |  |
| --- | --- | --- | --- | --- | --- |
|  | **N** | **% (95% CI)** | **N** | **% (95% CI)** | **Difference ^a^ (95% CI)** |
| **Pre-intervention period** | | | | | |
| 2011-12 | 8,441 | 32.5 (31.3, 33.7) | 8,562 | 38.4 (37.2, 39.6) | -5.9 (-7.6, -4.2) |
| 2012-13 | 8,568 | 36.3 (35.1, 37.5) | 8,552 | 42.5 (41.3, 43.8) | -6.3 (-8.0, -4.5) |
| 2013-14 | 8,654 | 43.3 (42.1, 44.6) | 8,537 | 49.0 (47.7, 50.2) | -5.7 (-7.4, -3.9) |
| **Intervention period** | | | | | |
| 2014-15 | 8,827 | 36.2 (35.1, 37.4) | 8,710 | 44.8 (43.6, 46.0) | -8.5 (-10.2, -6.9) |
| 2015-16 | 9,211 | 29.6 (28.5, 30.7) | 8,968 | 44.6 (43.4, 45.8) | -15.0 (-16.6, -13.4) |
| 2016-17 | 9,419 | 32.5 (31.4, 33.6) | 8,942 | 43.8 (42.6, 45.0) | -11.3 (-12.9, -9.6) |

Includes patients aged 5-12 years. Includes all influenza vaccinations at KPNC between September 1 and August 31 each year.

^a^ Percentage vaccinated for influenza at KPNC in the intervention site minus percentage in the comparison site. 95% CI accounts for clustering within households.

**Supplement Table 5. Unadjusted pre-intervention difference-in-differences**

|  | **2014-15** | | **2015-16** | | **2016-17** | |
| --- | --- | --- | --- | --- | --- | --- |
|  | Person-seasons | Estimate (95% CI) | Person-seasons | Estimate (95% CI) | Person-seasons | Estimate (95% CI) |
| **Medically attended acute respiratory infection per 1,000** | | | | | | |
| 0-4 years | 39,600 | -18.1 (-40.8, 4.5) | 39,923 | -12.5 (-34.2, 9.1) | 40,409 | -0.9 (-23.1, 21.2) |
| 5-12 years | 68,851 | -5.3 (-20.1, 9.5) | 69,493 | 3.9 (-8.8, 16.5) | 69,675 | -0.1 (-12.9, 12.8) |
| 13-17 years | 44,327 | 3.7 (-11.2, 18.7) | 44,633 | 5.9 (-6.7, 18.5) | 44,645 | 6.1 (-7.1, 19.3) |
| 18-64 years | 471,767 | -8.4 (-12.9, -4.0) | 480,303 | -2.8 (-6.7, 1.0) | 487,592 | -4.0 (-8.1, 0.1) |
| 65+ years | 104,623 | -13.2 (-23.2, -3.2) | 106,500 | -21.5 (-31.1, -11.9) | 108,060 | -13.0 (-23.2, -2.9) |
| **Laboratory-confirmed influenza positive among those tested per 100** | | | | | | |
| 0-4 years | 823 | -14.4 (-24.4, -4.5) | 765 | 10.0 (-2.7, 22.7) | 810 | -5.1 (-16.5, 6.3) |
| 5-12 years | 339 | -3.0 (-26.2, 20.1) | 323 | -4.3 (-29.5, 20.8) | 303 | -6.2 (-34.6, 22.2) |
| 13-17 years | 134 | -- | 136 | 23.5 (-15.3, 62.3) | 134 | -- |
| 18-64 years | 1,035 | 8.3 (-3.7, 20.3) | 1,007 | 13.1 (0.6, 25.7) | 1,164 | 0.7 (-10.3, 11.8) |
| 65+ years | 888 | -0.3 (-11.9, 11.3) | 837 | 4.1 (-8.3, 16.4) | 1,007 | 5.0 (-5.4, 15.3) |
| **Filled Oseltamivir prescription per 1,000** | | | | | | |
| 0-4 years | 39,600 | -0.2 (-5.0, 4.6) | 39,923 | -2.5 (-6.0, 0.9) | 40,409 | 1.6 (-2.2, 5.4) |
| 5-12 years | 68,851 | -0.4 (-4.1, 3.3) | 69,493 | -3.5 (-5.5, -1.5) | 69,675 | -4.0 (-6.5, -1.6) |
| 13-17 years | 44,327 | 1.9 (-1.4, 5.3) | 44,633 | 0.6 (-2.1, 3.3) | 44,645 | -1.4 (-4.5, 1.7) |
| 18-64 years | 471,767 | 0.3 (-0.7, 1.3) | 480,303 | 0.3 (-0.4, 1.0) | 487,592 | -1.6 (-2.5, -0.7) |
| 65+ years | 104,623 | 0.7 (-1.9, 3.3) | 106,500 | 1.7 (-0.2, 3.6) | 108,060 | 0.8 (-1.6, 3.1) |
| **Influenza hospitalization per 1,000** | | | | | | |
| 0-4 years | 39,600 | -0.9 (-5.4, 3.6) | 39,923 | -0.4 (-3.8, 3.1) | 40,409 | -1.1 (-5.0, 2.7) |
| 5-12 years | 68,851 | 1.6 (-0.8, 3.9) | 69,493 | -0.6 (-2.2, 1.1) | 69,675 | -0.2 (-2.0, 1.6) |
| 13-17 years | 44,327 | -0.3 (-3.1, 2.5) | 44,633 | -1.5 (-3.7, 0.6) | 44,645 | -1.3 (-3.7, 1.0) |
| 18-64 years | 471,767 | 4.1 (2.5, 5.7) | 480,303 | 1.9 (0.8, 3.0) | 487,592 | 1.7 (0.6, 2.9) |
| 65+ years | 104,623 | 8.0 (2.0, 14.1) | 106,500 | 0.7 (-3.7, 5.2) | 108,060 | 3.0 (-1.7, 7.7) |

Person-seasons are the number of individuals in a given season and in all pre-intervention seasons that were included in each difference-in-difference analysis. Estimates in children 5-12 years measure total effects, and estimates in other age groups measure indirect effects. Analyses were restricted to influenza season defined based the percentage of medical visits for influenza-like illness in California as reported by the California Department of Public Health. Influenza season started when there were at least 2 consecutive weeks in which the percentage of medical visits for influenza-like illness exceeded 2%, and the season ended when there were at least two consecutive weeks in which the percentage was less than or equal to 2%. Difference-in-difference estimates compare the difference in mean outcome in each district in an intervention year compared to the three pre-intervention years (2011-2013). Difference-in-difference parameters remove any time-invariant differences between groups (measured or unmeasured). Parameters were estimated using a generalized linear model without covariate adjustment due to data sparsity. Standard errors accounted for clustering at the household level. Estimates are not shown for some outcomes and ages due to sparse data.

**Supplement Table 6. Adjusted pre-intervention difference-in-differences**

|  | **2014-15** | | **2015-16** | | **2016-17** | |
| --- | --- | --- | --- | --- | --- | --- |
|  | Person-seasons | Estimate (95% CI) | Person-seasons | Estimate (95% CI) | Person-seasons | Estimate (95% CI) |
| **Medically attended acute respiratory infection per 1,000** | | | | | | |
| 0-4 years | 39,600 | -13.5 (-36.1, 9.1) | 39,923 | -7.4 (-28.9, 14.2) | 40,409 | 4.0 (-18.1, 26.0) |
| 5-12 years | 68,851 | -1.5 (-16.3, 13.2) | 69,493 | 7.0 (-5.6, 19.6) | 69,675 | 2.9 (-9.9, 15.7) |
| 13-17 years | 44,327 | 4.7 (-10.3, 19.6) | 44,633 | 6.8 (-5.7, 19.3) | 44,645 | 7.0 (-6.2, 20.1) |
| 18-64 years | 471,767 | -7.7 (-12.1, -3.3) | 480,303 | -1.6 (-5.4, 2.3) | 487,592 | -2.0 (-6.1, 2.1) |
| 65+ years | 104,623 | -12.3 (-22.3, -2.3) | 106,500 | -20.9 (-30.4, -11.4) | 108,060 | -12.3 (-22.4, -2.2) |
| **Laboratory-confirmed influenza positive among those tested per 100** | | | | | | |
| 0-4 years | 823 | -- | 765 | -- | 810 | -- |
| 5-12 years | 339 | -- | 323 | -- | 303 | -- |
| 13-17 years | 134 | -- | 136 | -- | 134 | -- |
| 18-64 years | 1,035 | 8.9 (-3.0, 20.8) | 1,007 | -- | 1,164 | 0.4 (-10.6, 11.5) |
| 65+ years | 888 | -- | 837 | -- | 1,007 | 5.1 (-5.2, 15.4) |
| **Filled Oseltamivir prescription per 1,000** | | | | | | |
| 0-4 years | 39,600 | -0.1 (-5.0, 4.7) | 39,923 | -- | 40,409 | -- |
| 5-12 years | 68,851 | -0.3 (-4.0, 3.4) | 69,493 | -- | 69,675 | -4.0 (-6.4, -1.5) |
| 13-17 years | 44,327 | 2.0 (-1.4, 5.3) | 44,633 | -- | 44,645 | -1.4 (-4.5, 1.7) |
| 18-64 years | 471,767 | 0.3 (-0.7, 1.3) | 480,303 | 0.3 (-0.4, 1.1) | 487,592 | -1.5 (-2.4, -0.6) |
| 65+ years | 104,623 | 0.7 (-1.8, 3.3) | 106,500 | 1.7 (-0.2, 3.7) | 108,060 | 0.8 (-1.6, 3.2) |
| **Influenza hospitalization per 1,000** | | | | | | |
| 0-4 years | 39,600 | -0.6 (-5.1, 3.9) | 39,923 | -- | 40,409 | -0.7 (-4.5, 3.1) |
| 5-12 years | 68,851 | 1.9 (-0.5, 4.2) | 69,493 | -- | 69,675 | -- |
| 13-17 years | 44,327 | -- | 44,633 | -- | 44,645 | -- |
| 18-64 years | 471,767 | 4.3 (2.7, 5.9) | 480,303 | 2.1 (1.1, 3.2) | 487,592 | 2.1 (0.9, 3.3) |
| 65+ years | 104,623 | 7.9 (1.9, 14.0) | 106,500 | 0.6 (-3.9, 5.1) | 108,060 | 2.8 (-1.9, 7.5) |

Person-seasons are the number of individuals in a given season and in all pre-intervention seasons that were included in each difference-in-difference analysis. Estimates in children 5-12 years measure total effects, and estimates in other age groups measure indirect effects. Analyses were restricted to influenza season defined based the percentage of medical visits for influenza-like illness in California as reported by the California Department of Public Health. Influenza season started when there were at least 2 consecutive weeks in which the percentage of medical visits for influenza-like illness exceeded 2%, and the season ended when there were at least two consecutive weeks in which the percentage was less than or equal to 2%. Difference-in-difference estimates compare the difference in mean outcome in each district in an intervention year compared to the three pre-intervention years (2011-2013). Difference-in-difference parameters remove any time-invariant differences between groups (measured or unmeasured). Parameters were estimated using a generalized linear model without covariate adjustment due to data sparsity. Standard errors accounted for clustering at the household level. Adjusted DID estimates were not available for some outcomes due to data sparsity.

**Supplement Table 7. Unadjusted pre-intervention difference-in-differences among individuals not vaccinated for influenza at KPNC**

|  | **2014-15** | | **2015-16** | | **2016-17** | |
| --- | --- | --- | --- | --- | --- | --- |
|  | Person-seasons | Estimate (95% CI) | Person-seasons | Estimate (95% CI) | Person-seasons | Estimate (95% CI) |
| **Medically attended acute respiratory infection per 1,000** | | | | | | |
| 0-4 years | 15,231 | 11.2 (-25.3, 47.7) | 15,485 | 8.0 (-24.7, 40.7) | 15,355 | 17.0 (-16.7, 50.7) |
| 5-12 years | 41,061 | 0.8 (-17.8, 19.3) | 42,075 | 11.8 (-3.1, 26.8) | 42,010 | -3.9 (-19.2, 11.5) |
| 13-17 years | 30,115 | 4.0 (-12.6, 20.6) | 30,204 | -3.6 (-17.5, 10.3) | 30,086 | -2.8 (-17.3, 11.7) |
| 18-64 years | 328,927 | -5.4 (-10.5, -0.3) | 336,187 | -3.3 (-7.6, 1.1) | 340,684 | -4.0 (-8.7, 0.6) |
| 65+ years | 34,052 | -20.7 (-37.4, -4.0) | 34,874 | -26.8 (-41.9, -11.6) | 35,534 | -23.0 (-39.2, -6.7) |
| **Laboratory-confirmed influenza positive among those tested per 100** | | | | | | |
| 0-4 years | 261 | -- | 249 | -- | 253 | -- |
| 5-12 years | 201 | -- | 199 | -12.3 (-44.8, 20.1) | 189 | -- |
| 13-17 years | 82 | -- | 79 | -- | 80 | -- |
| 18-64 years | 547 | 6.0 (-12.3, 24.3) | 561 | 24.7 (6.5, 42.9) | 650 | 4.6 (-11.1, 20.2) |
| 65+ years | 229 | -- | 209 | -- | 278 | 7.1 (-15.4, 29.6) |
| **Filled Oseltamivir prescription per 1,000** | | | | | | |
| 0-4 years | 15,231 | 1.9 (-6.6, 10.3) | 15,485 | -- | 15,355 | -1.3 (-8.8, 6.3) |
| 5-12 years | 41,061 | -2.5 (-7.1, 2.1) | 42,075 | -4.2 (-6.9, -1.5) | 42,010 | -5.5 (-8.8, -2.2) |
| 13-17 years | 30,115 | 2.8 (-0.9, 6.5) | 30,204 | 2.0 (-1.1, 5.1) | 30,086 | -1.4 (-5.1, 2.3) |
| 18-64 years | 328,927 | -0.1 (-1.2, 1.0) | 336,187 | 0.6 (-0.3, 1.4) | 340,684 | -1.9 (-2.9, -0.8) |
| 65+ years | 34,052 | -0.4 (-4.4, 3.5) | 34,874 | -0.1 (-3.2, 3.1) | 35,534 | 2.6 (-1.4, 6.5) |
| **Influenza hospitalization per 1,000** | | | | | | |
| 0-4 years | 15,231 | -2.2 (-8.7, 4.3) | 15,485 | -- | 15,355 | -- |
| 5-12 years | 41,061 | 1.2 (-1.4, 3.8) | 42,075 | -- | 42,010 | 0.6 (-1.6, 2.7) |
| 13-17 years | 30,115 | -- | 30,204 | -- | 30,086 | -- |
| 18-64 years | 328,927 | 3.2 (1.6, 4.7) | 336,187 | 1.1 (0.1, 2.2) | 340,684 | 1.1 (0.0, 2.2) |
| 65+ years | 34,052 | 11.8 (2.2, 21.4) | 34,874 | 0.2 (-6.5, 6.8) | 35,534 | 2.9 (-4.1, 9.8) |

Person-seasons are the number of individuals in a given season and in all pre-intervention seasons that were included in each difference-in-difference analysis. Estimates in children 5-12 years measure total effects, and estimates in other age groups measure indirect effects. Analyses were restricted to influenza season defined based the percentage of medical visits for influenza-like illness in California as reported by the California Department of Public Health. Influenza season started when there were at least 2 consecutive weeks in which the percentage of medical visits for influenza-like illness exceeded 2%, and the season ended when there were at least two consecutive weeks in which the percentage was less than or equal to 2%. Difference-in-difference estimates compare the difference in mean outcome in each district in an intervention year compared to the three pre-intervention years (2011-2013). Difference-in-difference parameters remove any time-invariant differences between groups (measured or unmeasured). Parameters were estimated using a generalized linear model without covariate adjustment due to data sparsity. Standard errors accounted for clustering at the household level. Estimates are not shown for some outcomes and ages due to sparse data.

**Supplement Table 8. Unadjusted pre-intervention difference-in-differences among individuals vaccinated for influenza at KPNC**

|  | **2014-15** | | **2015-16** | | **2016-17** | |
| --- | --- | --- | --- | --- | --- | --- |
|  | Person-seasons | Estimate (95% CI) | Person-seasons | Estimate (95% CI) | Person-seasons | Estimate (95% CI) |
| **Medically attended acute respiratory infection per 1,000** | | | | | | |
| 0-4 years | 24,369 | -40.8 (-69.9, -11.8) | 24,438 | -20.8 (-49.1, 7.5) | 25,054 | -14.6 (-43.0, 13.8) |
| 5-12 years | 27,790 | -4.7 (-29.4, 19.9) | 27,418 | 2.8 (-19.8, 25.3) | 27,665 | 13.6 (-8.9, 36.0) |
| 13-17 years | 14,212 | 1.3 (-28.9, 31.4) | 14,429 | 22.1 (-3.4, 47.7) | 14,559 | 15.4 (-10.8, 41.7) |
| 18-64 years | 142,840 | -16.6 (-25.5, -7.6) | 144,116 | -3.8 (-11.8, 4.1) | 146,908 | -5.2 (-13.5, 3.1) |
| 65+ years | 70,571 | -9.4 (-22.1, 3.3) | 71,626 | -18.7 (-30.9, -6.5) | 72,526 | -9.2 (-22.0, 3.6) |
| **Laboratory-confirmed influenza positive among those tested per 100** | | | | | | |
| 0-4 years | 562 | -- | 516 | -- | 557 | -- |
| 5-12 years | 138 | -- | 124 | -- | 114 | -- |
| 13-17 years | 52 | -- | 57 | -- | 54 | -- |
| 18-64 years | 488 | 6.5 (-8.2, 21.3) | 446 | -- | 514 | -9.0 (-23.1, 5.1) |
| 65+ years | 659 | -1.4 (-14.3, 11.5) | 628 | 1.8 (-11.7, 15.2) | 729 | 2.5 (-8.5, 13.4) |
| **Filled Oseltamivir prescription per 1,000** | | | | | | |
| 0-4 years | 24,369 | -1.4 (-7.1, 4.3) | 24,438 | 0.0 (-3.8, 3.9) | 25,054 | 3.4 (-0.7, 7.5) |
| 5-12 years | 27,790 | 3.9 (-2.5, 10.3) | 27,418 | -- | 27,665 | -1.8 (-5.4, 1.8) |
| 13-17 years | 14,212 | 0.5 (-6.3, 7.3) | 14,429 | -2.0 (-7.2, 3.1) | 14,559 | -1.4 (-7.2, 4.4) |
| 18-64 years | 142,840 | 1.4 (-0.8, 3.6) | 144,116 | -0.4 (-1.8, 1.1) | 146,908 | -0.9 (-2.7, 0.8) |
| 65+ years | 70,571 | 1.3 (-2.0, 4.6) | 71,626 | 2.6 (0.1, 5.0) | 72,526 | -0.3 (-3.2, 2.7) |
| **Influenza hospitalization per 1,000** | | | | | | |
| 0-4 years | 24,369 | -0.3 (-6.4, 5.7) | 24,438 | 0.3 (-4.5, 5.0) | 25,054 | -0.5 (-5.6, 4.7) |
| 5-12 years | 27,790 | 2.5 (-2.0, 7.1) | 27,418 | -- | 27,665 | -- |
| 13-17 years | 14,212 | -4.4 (-10.9, 2.0) | 14,429 | -- | 14,559 | -- |
| 18-64 years | 142,840 | 6.0 (2.1, 9.8) | 144,116 | 2.7 (0.0, 5.3) | 146,908 | 2.4 (-0.4, 5.3) |
| 65+ years | 70,571 | 6.1 (-1.6, 13.9) | 71,626 | 0.7 (-5.1, 6.4) | 72,526 | 2.5 (-3.5, 8.5) |

Person-seasons are the number of individuals in a given season and in all pre-intervention seasons that were included in each difference-in-difference analysis. Analyses were restricted to influenza season defined based the percentage of medical visits for influenza-like illness in California as reported by the California Department of Public Health. Influenza season started when there were at least 2 consecutive weeks in which the percentage of medical visits for influenza-like illness exceeded 2%, and the season ended when there were at least two consecutive weeks in which the percentage was less than or equal to 2%. Difference-in-difference estimates compare the difference in mean outcome in each district in an intervention year compared to the three pre-intervention years (2011-2013). Difference-in-difference parameters remove any time-invariant differences between groups (measured or unmeasured). Parameters were estimated using a generalized linear model without covariate adjustment due to data sparsity. Standard errors accounted for clustering at the household level.

**Supplement Table 9. Unadjusted pre-intervention difference-in-differences for negative control outcomes**

|  | **2014-15** | | **2015-16** | | **2016-17** | |
| --- | --- | --- | --- | --- | --- | --- |
|  | Person-seasons | Estimate (95% CI) | Person-seasons | Estimate (95% CI) | Person-seasons | Estimate (95% CI) |
| **Medically attended diarrhea per 1,000 – individuals not vaccinated for influenza at Kaiser** | | | | | | |
| 0-4 years | 15,231 | 7.5 (-2.3, 17.3) | 15,485 | 7.4 (0.4, 14.3) | 15,355 | 2.2 (-5.7, 10.1) |
| 5-12 years | 41,061 | -3.8 (-7.3, -0.3) | 42,075 | -1.0 (-3.9, 1.9) | 42,010 | 2.4 (-0.4, 5.2) |
| 13-17 years | 30,115 | -0.2 (-3.6, 3.2) | 30,204 | -- | 30,086 | -1.5 (-5.0, 1.9) |
| 18-64 years | 328,927 | 0.0 (-1.4, 1.5) | 336,187 | 0.3 (-0.9, 1.4) | 340,684 | -0.1 (-1.2, 1.1) |
| 65+ years | 34,052 | -2.7 (-8.5, 3.1) | 34,874 | 0.3 (-4.3, 4.9) | 35,534 | -1.5 (-6.5, 3.4) |
| **Medically attended gastrointestinal illness per 1,000 – individuals not vaccinated for influenza at Kaiser** | | | | | | |
| 0-4 years | 15,231 | 14.3 (1.5, 27.2) | 15,485 | -- | 15,355 | -0.5 (-10.4, 9.3) |
| 5-12 years | 41,061 | -0.9 (-6.3, 4.6) | 42,075 | -0.6 (-4.8, 3.7) | 42,010 | -1.1 (-5.3, 3.1) |
| 13-17 years | 30,115 | -1.3 (-5.9, 3.3) | 30,204 | -- | 30,086 | -- |
| 18-64 years | 328,927 | 0.4 (-0.9, 1.7) | 336,187 | 0.4 (-0.6, 1.4) | 340,684 | -0.5 (-1.6, 0.5) |
| 65+ years | 34,052 | -- | 34,874 | -- | 35,534 | 1.5 (-1.6, 4.6) |
| **Medically attended diarrhea per 1,000 – individuals vaccinated for influenza at Kaiser** | | | | | | |
| 0-4 years | 24,369 | -3.4 (-13.3, 6.6) | 24,438 | 1.4 (-6.3, 9.0) | 25,054 | 7.0 (-0.9, 14.8) |
| 5-12 years | 27,790 | -0.4 (-6.0, 5.1) | 27,418 | 2.8 (-1.2, 6.9) | 27,665 | 1.9 (-2.6, 6.3) |
| 13-17 years | 14,212 | 4.2 (-2.9, 11.3) | 14,429 | 5.0 (-1.0, 11.0) | 14,559 | -- |
| 18-64 years | 142,840 | -0.7 (-3.3, 2.0) | 144,116 | -0.2 (-2.4, 1.9) | 146,908 | -0.3 (-2.6, 1.9) |
| 65+ years | 70,571 | 1.2 (-3.4, 5.9) | 71,626 | 2.1 (-1.6, 5.9) | 72,526 | 1.3 (-2.6, 5.3) |
| **Medically attended gastrointestinal illness per 1,000 – individuals vaccinated for influenza at Kaiser** | | | | | | |
| 0-4 years | 24,369 | -3.1 (-14.7, 8.6) | 24,438 | -4.3 (-12.7, 4.0) | 25,054 | -2.2 (-11.4, 7.1) |
| 5-12 years | 27,790 | 2.6 (-5.1, 10.3) | 27,418 | -5.9 (-11.8, 0.0) | 27,665 | -7.2 (-13.9, -0.6) |
| 13-17 years | 14,212 | 7.0 (-2.2, 16.1) | 14,429 | -3.3 (-10.1, 3.5) | 14,559 | 1.4 (-5.5, 8.2) |
| 18-64 years | 142,840 | 2.1 (-0.1, 4.4) | 144,116 | -0.3 (-2.2, 1.5) | 146,908 | -0.2 (-2.1, 1.7) |
| 65+ years | 70,571 | 0.6 (-2.3, 3.5) | 71,626 | 0.5 (-1.8, 2.8) | 72,526 | -2.0 (-4.4, 0.4) |

Person-seasons are the number of individuals in a given season and in all pre-intervention seasons that were included in each difference-in-difference analysis. Analyses were restricted to influenza season defined based the percentage of medical visits for influenza-like illness in California as reported by the California Department of Public Health. Influenza season started when there were at least 2 consecutive weeks in which the percentage of medical visits for influenza-like illness exceeded 2%, and the season ended when there were at least two consecutive weeks in which the percentage was less than or equal to 2%. Difference-in-difference estimates compare the difference in mean outcome in each district in an intervention year compared to the three pre-intervention years (2011-2013). Difference-in-difference parameters remove any time-invariant differences between groups (measured or unmeasured). Parameters were estimated using a generalized linear model without covariate adjustment due to data sparsity. Standard errors accounted for clustering at the household level.

**Supplement Table 10. Unadjusted pre-intervention difference-in-differences for negative control time period analysis**

|  | **2014-15** | | **2015-16** | | **2016-17** | |
| --- | --- | --- | --- | --- | --- | --- |
|  | Person-seasons | Estimate (95% CI) | Person-seasons | Estimate (95% CI) | Person-seasons | Estimate (95% CI) |
| **Medically attended acute respiratory infection per 1,000** | | | | | | |
| 0-4 years | 15,231 | 9.1 (-16.9, 35.2) | 15,485 | -12.8 (-42.2, 16.6) | 15,355 | 7.7 (-22.7, 38.2) |
| 5-12 years | 41,061 | 7.2 (-4.5, 19.0) | 42,075 | 6.7 (-7.3, 20.7) | 42,010 | -5.2 (-19.8, 9.4) |
| 13-17 years | 30,115 | 6.4 (-4.9, 17.8) | 30,204 | 13.6 (-0.8, 28.0) | 30,086 | 11.8 (-3.1, 26.7) |
| 18-64 years | 328,927 | 2.1 (-1.2, 5.3) | 336,187 | -8.1 (-12.2, -3.9) | 340,684 | -1.7 (-6.0, 2.6) |
| 65+ years | 34,052 | -3.1 (-15.6, 9.5) | 34,874 | 1.5 (-13.7, 16.7) | 35,534 | 0.3 (-14.8, 15.3) |
| **Influenza hospitalization per 1,000** | | | | | | |
| 0-4 years | 15,231 | 1.2 (-3.3, 5.7) | 15,485 | -- | 15,355 | -- |
| 5-12 years | 41,061 | 0.5 (-1.6, 2.7) | 42,075 | -- | 42,010 | -0.9 (-3.2, 1.4) |
| 13-17 years | 30,115 | -- | 30,204 | -- | 30,086 | -- |
| 18-64 years | 328,927 | 1.6 (0.4, 2.8) | 336,187 | 0.1 (-1.2, 1.4) | 340,684 | 0.8 (-0.4, 2.1) |
| 65+ years | 34,052 | 7.8 (-0.2, 15.9) | 34,874 | -1.1 (-9.1, 7.0) | 35,534 | 1.2 (-6.7, 9.0) |

Person-seasons are the number of individuals in a given season and in all pre-intervention seasons that were included in each difference-in-difference analysis. Analyses were restricted to the period outside of influenza season defined based the percentage of medical visits for influenza-like illness in California as reported by the California Department of Public Health. Influenza season started when there were at least 2 consecutive weeks in which the percentage of medical visits for influenza-like illness exceeded 2%, and the season ended when there were at least two consecutive weeks in which the percentage was less than or equal to 2%. Difference-in-difference estimates compare the difference in mean outcome in each district in an intervention year compared to the three pre-intervention years (2011-2013). Difference-in-difference parameters remove any time-invariant differences between groups (measured or unmeasured). Parameters were estimated using a generalized linear model without covariate adjustment due to data sparsity. Standard errors accounted for clustering at the household level.
